# Pattern of variation in DNA methylation during pregnancy among mothers who delivered preterm in the GARBH-Ini cohort

**DOI:** 10.1101/2021.09.02.21262698

**Authors:** Jagyashila Das, Indranil Bagchi, Shekhar Ghosh, Nitya Wadhwa, Uma Chadramouli Nachu, Ramachandran Thiruvengadam, Pallavi Kshetrapal, Shinjini Bhatnagar, Partha P. Majumder, Arindam Maitra, GARBH-Ini Team

## Abstract

**Background:** DNA methylation (DNAm) may play an important role in birth outcomes.

**Material and Methods:** Genome wide DNAm was analysed in peripheral blood DNA of women at multiple time points during gestation. A novel empirical method was used to identify CpG sites with high temporal variance in methylation associating with preterm birth.

**Results:** High variability at 1296 CpG sites from the promoter regions of 1197 genes significantly associated with PTB. These genes belonged to pathways involved in signalling by platelet derived growth factor, platelet homeostasis, collagen degradation, extracellular matrix and circadian clock.

**Conclusions:** The findings provide novel information which might help in development of predictive biomarkers of preterm birth outcome.

## Introduction

Preterm birth (PTB) is any live birth before 37 completed weeks of gestation [1]. It is the leading cause of neonatal morbidity. Individuals born preterm face greater risk of delayed development of cognitive and motor skills, sensory deficits, learning disabilities and have higher risk of cerebral palsy, respiratory illness, immunodeficiency and other late onset diseases, etc. [2,3]. In India, 3.6 million of 27 million babies are born preterm every year [4]. PTB is a syndrome attributable to multiple etiologies [5,6]. Epidemiological studies suggest increased risk of PTB can be attributed to nutritional deficiencies, stress, and other factors such as smoking, maternal infection (bacterial vaginosis/intra-amniotic infection), and race [7]. These factors can modulate cellular functioning primarily via DNA methylation [8–10].

Biochemical and molecular signatures in peripheral blood provide a unique potential window into the health and phenotype of an individual. Longitudinal epigenomic profiling of peripheral blood of pregnant women across all three trimesters of pregnancy and at delivery is expected to provide an improved understanding of the physiological changes that take place during gestation, in relation to birth outcome, including PTB [11]. Currently, limited data are available on the role of maternal DNA methylation alterations in pregnancy outcomes; available in only African-American population [12,13]. Blood samples for these studies were collected either at labor or shortly after delivery and hence do not capture the longitudinal changes that take place during the course of pregnancy. A detailed review on existing studies of maternal DNAm during pregnancy discusses the existing limitations in the field [14].

A multidisciplinary Group for Advanced Research on Birth-outcomes -DBT India Initiative (GARBH-Ini) program, was established in 2014 to identify clinical, epidemiologic, genomic, epigenomic, proteomic, metabolic and microbial correlates of PTB [15]. A cohort (named the GARBH-Ini cohort) of pregnant women was established as part of this programme with the primary objective of generating a risk-prediction algorithm for preterm birth based on multidimensional risk factors assessed during pregnancy, using a time series approach. We have undertaken the present study, on the GARBH-Ini platform, to test the hypothesis that increased epigenomic variation in maternal blood DNA during pregnancy results in preterm birth. In particular, we hypothesized that instability in the extent of promoter DNA methylation during pregnancy of key genes that regulate the period of gestation will be associated with preterm birth. The instability in methylation can be measured by the variance in the level of methylation during the course of pregnancy.

Thus, we posit that women who delivered preterm will exhibit greater temporal variance in DNA methylation during gestation, including at time of delivery, compared to those who delivered at term.

## Methods

### Study Participants

GARBH-Ini cohort was established at the district hospital in Gurugram, Haryana, India. Women are enrolled within 20 weeks of conception as assessed by ultrasonography and are followed up at least once during each trimester during the antenatal period until delivery and once within 6 months of post-partum. Detailed information is collected from the participants on their socio-demography, environmental and clinical details, nutritional parameters. Serial ultrasound scans are done across pregnancy for uterine, fetal and placental morphology and biospecimens are collected from each woman during the course of her pregnancy at defined intervals. The profile, detailed objectives and methodology of this cohort, have been published earlier [15].

This study was designed as case-control study nested in GARBH-Ini cohort. Approvals from Institutional Ethics Committees of Gurugram Civil Hospital, Translational Health Science and Technology Institute, National Institute of Biomedical Genomics and the collaborating hospitals were obtained. Each woman recruited into this study provided prior informed voluntary consent for participation. The study population from which the women who delivered preterm -- a live birth less than 37 completed weeks of gestation -- and at term were sampled included those who gave birth to singleton baby by spontaneous delivery without congenital abnormalities or medical complications during pregnancy and had visited the clinical site <14 weeks, 18-20, 26-28 and at delivery to contribute, with informed consent, biospecimens for analysis. Time of sample collection for each study participant is provided in supplementary table 2. Mothers with concomitant complexities like intrauterine growth restriction (IUGR), uterine infection, preeclampsia, any medical or surgical illness, e,g., HIV, malignancy, immunodeficiency or a history of long-term intake of immunomodulators and steroids were excluded. A total of 17 pairs of mothers, who delivered at term and preterm, matched for month of delivery and parity, were selected for this study based on the criteria described above.

### Sample collection and DNA methylation assay

Maternal peripheral blood samples were collected in EDTA vacutainers at 11-14 weeks, 18-20 weeks, 26-28 weeks of gestation and at the time of delivery for DNA extraction from each study participant. DNA was isolated using QIAamp blood midi kit (Qiagen) and quantified in Qubit fluorimeter (Thermo). Bisulphite conversion of genomic DNA was carried out using the EZ DNA methylation kit (Zymo Research) and used for Infinium MethylationEPIC BeadChip (Illumina) according to the manufacturer’s guidelines. The processed chips were scanned using iScan reader (Illumina) and raw data were analyzed using the methylation module under the GenomeStudio software (Illumina) version 2011.1.

DNA methylation levels for each probe was converted from fluorescence intensities of methylated (M) and unmethylated (U) alleles to methylation level, ranging from 0 (no methylation) to 1 (completely methylated), given by β=M/(M+U+100) [16].

Probes with <3 beads in at least 5% of samples, along with 2991 non-CpG probes, 81493 CpG sites which were identified by Illumina as SNPs, 49 multi-hit probes along with 2991 non-cpG probes, and 17844 probes on the sex chromosomes were removed using ChAMP package version 2.8.3 [17–19] in R version 3.6.3 (2020-02-29) [20].

Illumina EPIC methylation array uses two types of probes which have different dynamic ranges as well as a type 2 probe bias. Beta Mixture Quantile dilation (BMIQ) was used to normalise for the two types of probes used in the array [21]. Quantile normalisation (QN) was performed to equalize the distribution of probe intensities from each array [22,23]. Cell type proportion using the algorithm described previously for CD4+ T cells, CD8+ T cells, natural killer (NK) cells, B cells, monocytes, and granulocytes were estimated using champ.refbase command from the ChAMP analysis pipeline to account for the potential differences in DNA methylation (DNAm) that arise due to cell composition variability in whole blood [19,24].

We used Illumina Infinium EPIC manifest file in which annotation is based on hg19 reference sequence. We considered probes in promoter specific regions of the genome, i.e. from 200 to 1500 bases upstream of transcription start sites and 5’ untranslated regions (TSS200, TSS1500, 5’UTR) for analysis. We have used probes with β value ranging from 0.2 to 0.8 (corresponding M value ranging from -2 to 2) for subsequent analysis [25]. We proceeded with 26,107 probes for subsequent analysis.

### Statistical Method

We performed unpaired two-tailed t-tests to compare equality of mean values of the period of gestation (POG), maternal age at conception, and infant birth weight between the two groups of mothers. We tested whether sex of the child is associated with the status of the birth outcome using a chi-squared test.

To test the equality of temporal variances between the two groups over the four time points during gestation and at delivery at which biospecimens were collected, we adopted the following empirically-driven statistical approach.

For each mother and each promoter probe, we calculated V_ij_, variance of M _ij_^(1)^, M_ij_ ^(2)^, M_ij_ ^(3)^, M_ij_ ^(4)^, where M_ij_ ^(t)^ denotes the M value of the j^th^ probe(j=1,2, 103517) for the i^th^ mother (preterm group= 1,2,…17; term group= 18,19,….34). We then computed 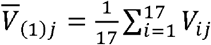 and 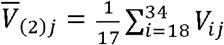, the mean values of temporal variances across mothers in each of the two groups, for the j^th^ probe; and, 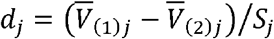, where 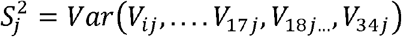. It may be noted that d_j_ is analogous to the t-statistic for testing equality of mean values.

In other words, d_j_ is the statistic to compare equality of temporal variances in methylation for the j^th^ probe. To test the statistical significance of the observed d_j_, we calculated an empirical sampling distribution of the d_j_-statistic by re-sampling M values with replacement for the j^th^ probe for the 34 mothers, using the spirit of bootstrapping. An observed value of d_j_ was declared to be significant at the 1% level if the value of the d_j_ belonged to the upper 1% tail of the empirical distribution.

We used the list of promoter probes for which the temporal variance between mothers who delivered at term and preterm were significantly different. We mapped these probes to promoter regions of genes. A brief overview of the workflow is represented in Supplementary figure 1 and supplementary table 1.

### Pathway Enrichment Analysis

The 1197 genes identified as mentioned above were analysed for enrichment of biological pathways. REACTOME [26] and KEGG [27] data resources were used in this analysis. ClueGO V2.5.1 plugins [28] and CluePedia V1.5.1 [29] and of Cytoscape (v. 3.7.0) [30] were used to analyse interrelations of terms and pathways in biological networks and visualise known interactions of genes in pathways from the data matrix, respectively. For the enrichment, a right-sided hyper-geometric test was used. The Kappa score threshold value was set to ≥0.4. Criteria to identify a pathway as significant were that the pathway (a) should have minimum of 10 genes and (b) the p-value corrected for multiple-testing using the Benjamini-Hochberg method should be <0.05.

## Results

### Participant details

The summary of demographic and other information of the participants are provided in Table 1, and Supplementary Figure 2. For the 17 pairs of mothers, who delivered preterm (mean POG 33.1±2.8 weeks) and at term (mean POG 39.2±1.2 weeks), there were no significant difference in mean maternal age (t = -1.5765, df = 32, p= 0.1248) or any association of sex of the child with status of birth (χ2 = 1.91, df = 1, p = 0.1671). As expected, there was a significant difference in mean weight of a child (mean weight PTB= 1.92kg, mean weight term = 2.77kg) at birth (t = 5.0066, df = 32, p= 1.949 ×10-05).

**Table 1:** Summary of information of mothers included in the study.

| Variable | Statistic |  | Mothers delivering |  |
| --- | --- | --- | --- | --- |
|  |  |  | Preterm (n=17) | Term (n=17) |
| Maternal age (years) | Minimum |  | 19 | 19 |
|  | Maximum |  | 27 | 29 |
|  | Mean |  | 21.3 | 22.8 |
|  | SD |  | 2.4 | 3.2 |
| Period of gestation* (weeks) | Minimum |  | 27.1 | 37 |
|  | Maximum |  | 35.8 | 41.6 |
|  | Mean |  | 33.1 | 39.2 |
|  | SD |  | 2.8 | 1.2 |
| Weight of child at birth* (kg) | Minimum |  | 1.04 | 2.29 |
|  | Maximum |  | 3.22 | 3.34 |
|  | Mean |  | 1.92 | 2.77 |
|  | SD |  | 0.62 | 0.32 |
| Proportion of female: male children at birth |  |  | 5:12 | 10:7 |
| BMI category | Underweight |  | 7 (41.18%) | 3 (17.65%) |
|  | Normal |  | 9 (52.94%) | 11 (64.71%) |
|  | Overweight |  | 1 (5.88%) | 2 (11.76%) |
|  | Obese |  | - | 1 (5.88%) |
| History of vaginal discharge | Present |  | 1 (5.88%) | 1 (5.88%) |
|  | Absent |  | 16 (94.12%) | 16 (94.12%) |
| History of bleeding per vagina | Present |  | 2 (11.76%) | - |
|  | Absent |  | 15 (88.24%) | 17 (100%) |
| Socioeconomic status | Upper Middle Class |  | 3 (17.65%) | 3 (17.65%) |
|  | Lower Middle Class |  | 10 (58.82%) | 3 (17.65%) |
|  | Upper Lower Middle Class |  | 4 (23.53%) | 10 (58.82%) |
|  | Lower Class |  | - | 1 (5.88%) |
| Toilet usage | Flush/ pour flush toilet |  | 16 (94.12%) | 17 (100.00%) |
|  | Bucket latrine |  | 1 (5.88%) | - |
| Laboratory characteristics | POG at sampling | Enrolment | 11w3d (8w1d,12w6d) | 10w3d (8w1d,12w4d) |
|  |  | Visit 2 | 18w4d (18w4d,19w4d) | 18w4d (18w2d,19w5d) |
|  |  | Visit 3 | 26w2d (26w1d,26w3d) | 26w2d (26w1d,26w3d) |

\*Difference is significant ( $p < 0.05$ ) between term and preterm delivering mothers.

### DNA methylation analysis

Of the 219,163 probes located in the promoter regions of genes, CpG sites with M values ≤+2 and ≥-2 were not considered in all samples. This is to reduce the number of non-variable sites to further reduce the statistical burden of multiple testing in the subsequent analyses, as used in previous studies [31,32]. Data on the 26,107 CpG sites that remained were analyzed further. We identified 1296 probes from 1197 genes with standardized difference in mean values of temporal variance of methylation between the two groups of mothers above the 99th percentile of the distribution of empirical standardized differences. Mean and SD values are provided in supplementary Table 3.

The ten probes with the largest d-statistics belonged to *UST, OSBPL3, CGNL1, TBC1D24, NFYC, ZNF165, BMP5, CKM, C5orf47, TNRC6B* genes (Figure 1). The probe with the largest effect size of 1.76 in our analysis was cg03366867 which belonged to the promoter of *UST*.

**Fig 1.**
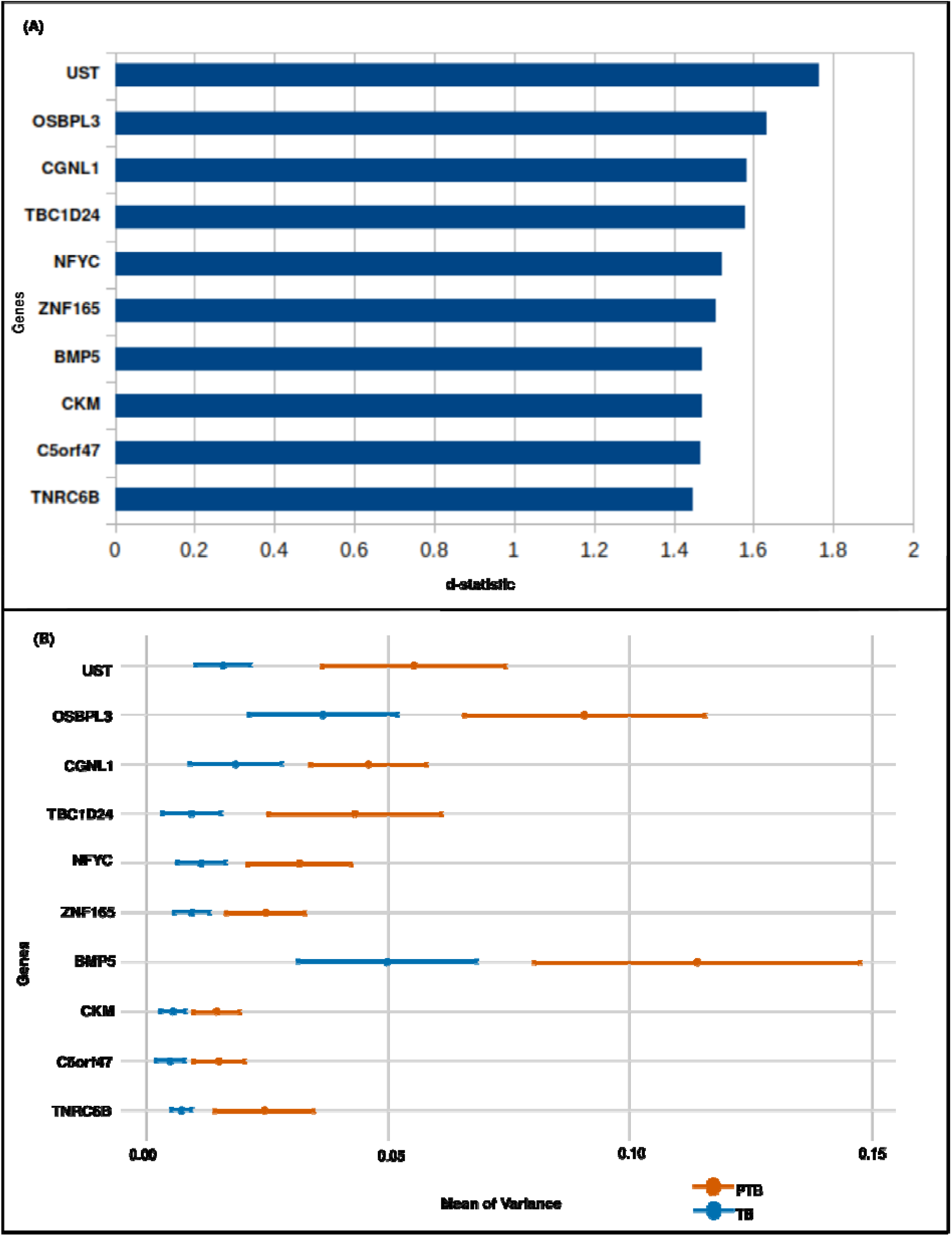
**(a)** Genes arranged in descending order of value of the d-statistic (**b)** Ten genes with the largest standardized mean difference in temporal variance of methylation between the mothers who delivered at term and preterm plotted with the mean of variance and confidence interval.

The four CpG sites from *UST* (cg03366867), *OSBPL3* (cg00891520), *CGNL1* (cg26185468), *TBC1D24* (cg08190844) with the highest temporal variation in PTB group are depicted in Figure 2.

**Fig 2.**
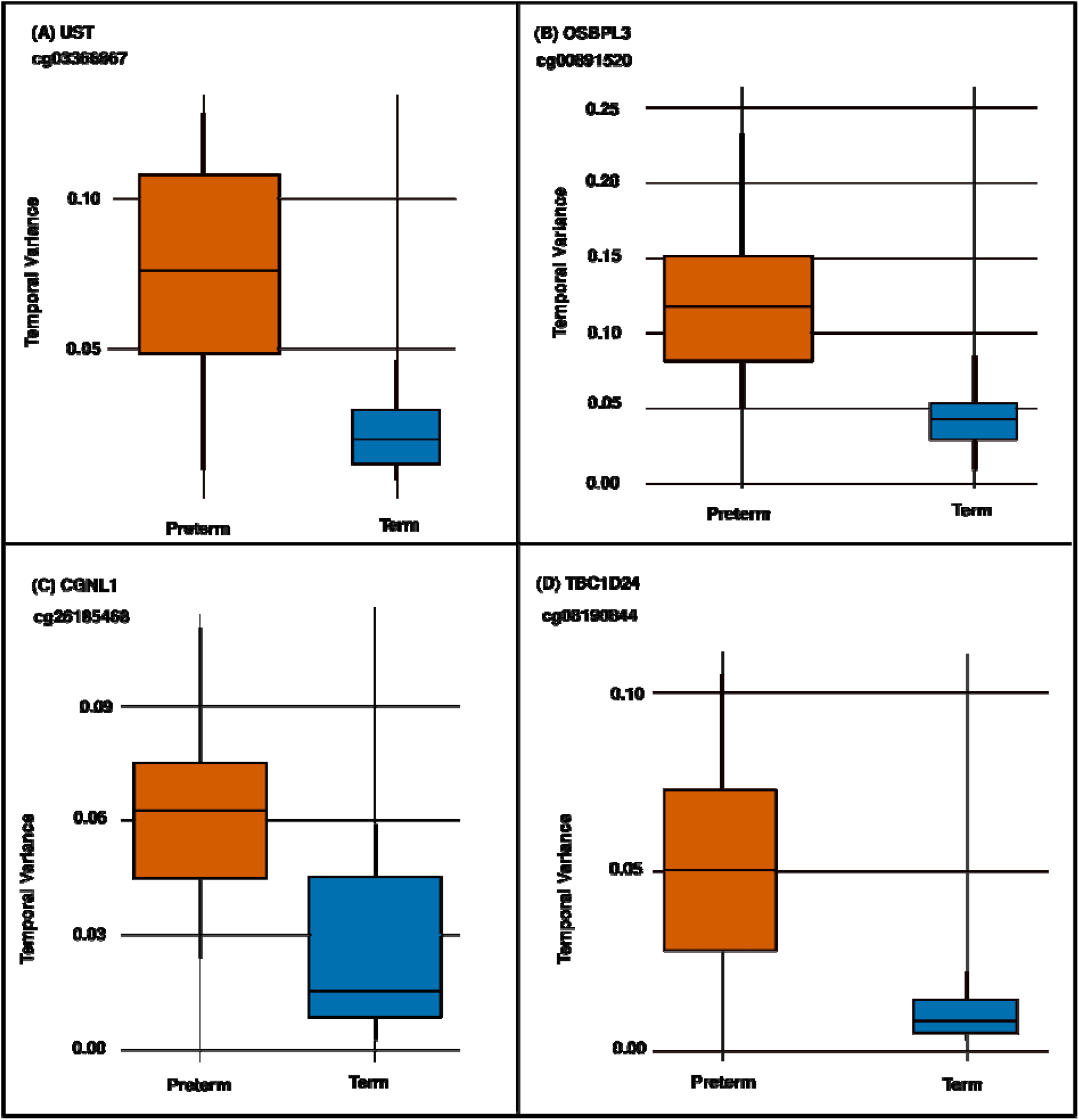
**(A-D)** Temporal variance of the probes with largest d-statistic in the x axis and status in the Y-axis.

### Pathway enrichment

Pathway analysis with the significant probes (1296 promoter probes from 1197 genes) that showed the highest standardised mean differences in temporal variance of DNA methylation between the two groups of mothers resulted in identification of twenty enriched pathways (BH corrected P <0.05). List of probes (1296) along with their corresponding d-statistics are provided in supplementary table 4. The relative abundance of the most significant pathways based on the BH corrected p-values are provided in Figure 3. The most significantly enriched pathway was signaling by platelet derived growth factor (*COL2A1, COL9A2, COL9A3, FURIN, GRB2, PLAT, RAPGEF1, SRC, STAT1, STAT3, STAT5B, THBS2*), followed by collagen degradation (*COL12A1, COL1A2, COL26A1, COL2A1, COL8A2, COL9A2, COL9A3, CTSB, CTSK, FURIN, MMP19*), platelet homeostasis (*ATP2B2, ATP2B4, FGR, GNAS, GNB1, GNG12, GNG4, GNG7, KCNMB1, ORAI1, PDE5A, PPP2R5A, PRKG2, SLC8A1, TRPC6*), circadian clock pathways (*AVP, CPT1A, CRY2, F7, NCOA1, NCOA2, NCOR1, NR3C1, PPP1CA, RAI1, UBB*). Extracellular matrix (ECM) proteoglycans and ECM-receptor interaction genes also exhibited higher temporal variation in PTB delivering mothers compared to those who delivered at term. The list of the 14 enriched pathways are provided in Table 2.

**Table 2:** Details of twenty significantly enriched pathways identified based on differential DNA methylation results.

| **GO Term** | **Term PValue**<br>**BH-**<br>**Corrected** | **%**<br>**Associated**<br>**Genes** | **No.of**<br>**genes** | **Associated**<br>**Genes** |
| --- | --- | --- | --- | --- |
| **Signaling by PDGF** | 0.01 | 20.69 | 12.00 | COL2A1,<br>COL9A2,<br>COL9A3,<br>FURIN, GRB2,<br>PLAT,<br>RAPGEF1, SRC,<br>STAT1, STAT3,<br>STAT5B, THBS2 |
| **Collagen degradation** | 0.03 | 17.19 | 11.00 | COL12A1,<br>COL1A2,<br>COL26A1,<br>COL2A1,<br>COL8A2,<br>COL9A2,<br>COL9A3, CTSB,<br>CTSK, FURIN,<br>MMP19 |
| **Platelet homeostasis** | 0.01 | 17.05 | 15.00 | ATP2B2,<br>ATP2B4, FGR,<br>GNAS, GNB1,<br>GNG12, GNG4,<br>GNG7,<br>KCNMB1,<br>ORAI1, PDE5A,<br>PPP2R5A,<br>PRKG2,<br>SLC8A1, TRPC6 |
| **Circadian Clock** | 0.05 | 15.71 | 11.00 | AVP, CPT1A,<br>CRY2, F7,<br>NCOA1,<br>NCOA2,<br>NCOR1, NR3C1,<br>PPP1CA, RAI1,<br>UBB |
| **ECM proteoglycans** | 0.05 | 14.47 | 11.00 | ACAN,<br>COL1A2,<br>COL2A1,<br>COL9A2,<br>COL9A3, DCN,<br>ITGA2B,<br>ITGA8, LRP4,<br>MATN4, TNXB |
| **Potassium Channels** | 0.04 | 14.14 | 14.00 | GNB1, GNG12,<br>GNG4, GNG7,<br>KCNA3,<br>KCNA4, |
|  |  |  |  | KCNA6,<br>KCNAB2,<br>KCNH2,<br>KCNJ10,<br>KCNJ8, KCNK3,<br>KCNMB1,<br>KCNQ3 |
| **ECM-receptor interaction** | 0.05 | 13.64 | 12.00 | COL1A2,<br>COL2A1,<br>COL9A2,<br>COL9A3,<br>GP1BA, GP6,<br>ITGA2B,<br>ITGA8, LAMC3,<br>SV2B, THBS2,<br>TNXB |
| **GABAergic synapse** | 0.05 | 13.48 | 12.00 | GABRG2,<br>GNB1, GNG12,<br>GNG4, GNG7,<br>PRKACB,<br>PRKCA,<br>SLC38A1,<br>SLC6A1,<br>SLC6A12, SRC,<br>TRAK2 |
| **Cargo recognition for**<br>**clathrin-mediated**<br>**endocytosis** | 0.05 | 13.21 | 14.00 | AP2B1, AP2M1,<br>AVP, CD3D,<br>CHRM2, EPGN,<br>EPN2, EPS15,<br>GRB2, NECAP2,<br>SLC2A8,<br>STON1-GTF2A1L,<br>SYT2, UBB |
| **Dopaminergic synapse** | 0.05 | 12.12 | 16.00 | CREB3L1,<br>CREB5, GNAL,<br>GNAS, GNB1,<br>GNG12, GNG4,<br>GNG7,<br>MAPK10,<br>MAPK12,<br>PPP1CA,<br>PPP2R2B,<br>PPP2R5A,<br>PRKACB,<br>PRKCA, SCN1A |
| **Hippo signaling pathway** | 0.04 | 11.69 | 18.00 | BMP5, BMP7,<br>BMP8B,<br>BMPR1A,<br>BMPR1B,<br>CTNNA2,<br>FRMD1, |
|  |  |  |  | FRMD6,<br>PARD6G,<br>PPP1CA,<br>PPP2R2B, TCF7,<br>TEAD1, TEAD3,<br>TEAD4,<br>TP53BP2, TP73,<br>YWHAG |
| **Adrenergic signaling in**<br>**cardiomyocytes** | 0.04 | 11.41 | 17.00 | ATP2B2,<br>ATP2B4,<br>CACNB2,<br>CACNG4,<br>CREB3L1,<br>CREB5, GNAS,<br>KCNE1,<br>MAPK12,<br>PPP1CA,<br>PPP2R2B,<br>PPP2R5A,<br>PRKACB,<br>PRKCA,<br>RAPGEF4,<br>SLC8A1,<br>TNNT2 |
| **Focal adhesion** | 0.05 | 10.55 | 21.00 | COL1A2,<br>COL2A1,<br>COL9A2,<br>COL9A3, GRB2,<br>ITGA2B,<br>ITGA8, LAMC3,<br>MAPK10,<br>MYLK, PAK5,<br>PAK6, PARVG,<br>PGF, PPP1CA,<br>PRKCA, PXN,<br>RAPGEF1, SRC,<br>THBS2, TNXB |
| **Neuronal System** | 0.01 | 10.24 | 42.00 | AP2B1, AP2M1,<br>APBA2,<br>ARHGEF7,<br>BCHE,<br>CACNB2,<br>CACNG4,<br>CHRNA1,<br>DLGAP1,<br>GABRG2,<br>GNAL, GNB1,<br>GNG12, GNG4,<br>GNG7, GRIK4,<br>KCNA3,<br>KCNA4,<br>KCNA6, |
|  |  |  |  | KCNAB2,<br>KCNH2,<br>KCNJ10,<br>KCNJ8, KCNK3,<br>KCNMB1,<br>KCNQ3,<br>NRXN2,<br>PPFIA4,<br>PPFIBP1,<br>PPFIBP2,<br>PRKACB,<br>PRKAR1B,<br>PRKCA,<br>RPS6KA1,<br>SHANK1,<br>SHANK2,<br>SLC38A1,<br>SLC6A1,<br>SLC6A12, SRC,<br>SYT2, TUBB3 |

**Fig 3.**
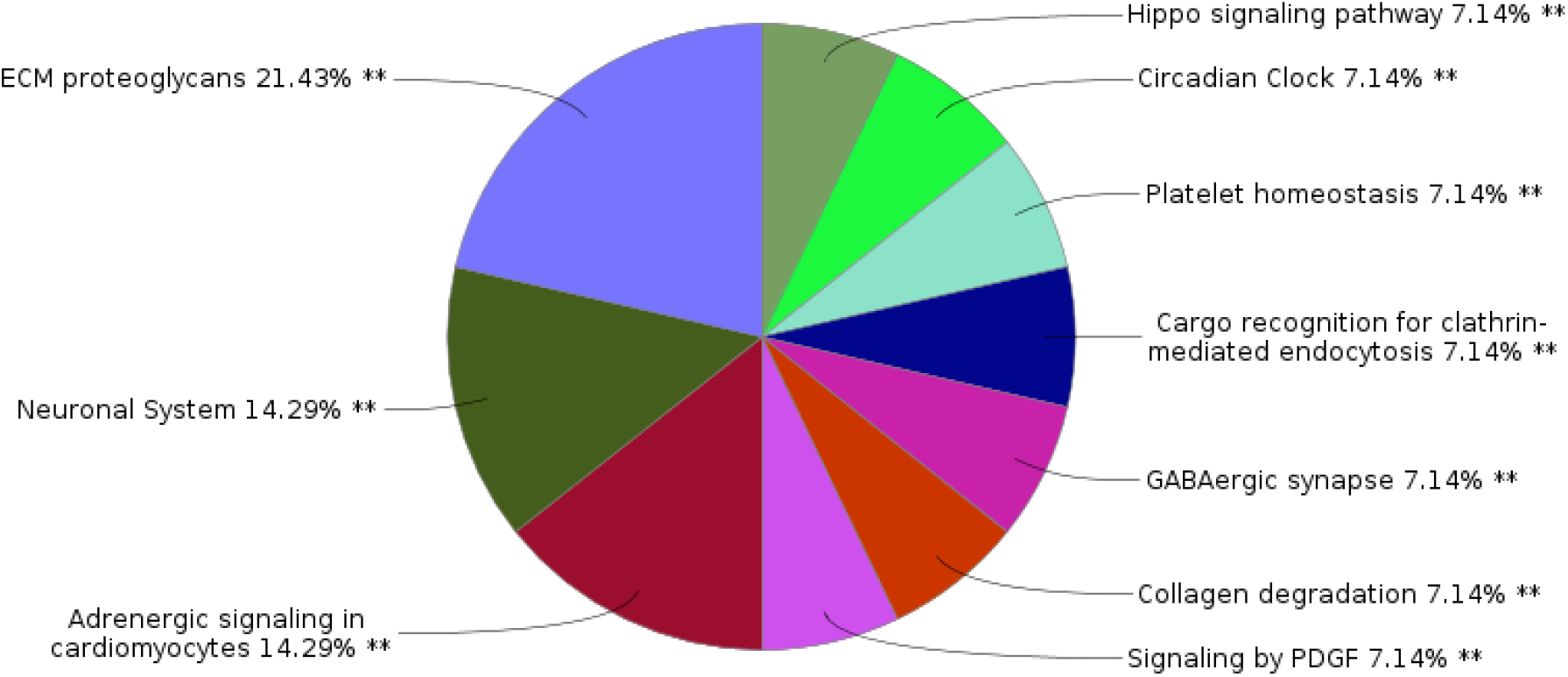
Percentage of terms per group, most significant pathways with BH corrected p-value (<0.05)

## Discussion

Pregnancy is a synchronized cascade of multiple physiological events triggered by changes in gene expression [32]. In view of the emerging perception that adverse pregnancy outcomes like PTB might result from dysregulation of this cascade, we hypothesized that dysregulation in DNA methylation changes during pregnancy may be a good predictor of PTB. Some studies have been initiated on maternal blood transcriptomic alterations during healthy pregnancy and adverse events associated with pregnancy [33]. However, the inherent problems associated with the transient nature of the alterations in mRNA levels in whole blood and the impact on stability of these mRNA by non-physiological factors related to extraction and storage, might limit the ability of such investigations. On the other hand, DNA methylation, used in our study, is expected to be unaffected by the above problems [34,35].

In our report, we have used a novel statistical method of analyzing temporal variation in DNA methylation between two groups of mothers with contrasting birth outcomes which allows for robust identification of large effects even in modest sample sizes. Methods to compare epigenome wide DNA methylation levels between these groups at individual time points may not have revealed the temporal differences between these groups of mothers during the course of pregnancy. We have identified those loci which exhibit larger variance over pregnancy in mothers delivering preterm birth giving compared to those who delivered at term. We view our findings as exploratory and recognize the importance of validating these in an independent set of samples. Nutrition and environmental exposures are important factors that may change the epigenetic pattern and it would be interesting to evaluate whether these findings hold true for women with different nutritional status as assessed by BMI and with variations in the environmental exposures like ambient air pollution.

Changes in DNA methylation of the promoter CpG sites might lead to altered gene expression[36,37]. We identified promoter CpG sites in 1197 genes with the highest difference in mean levels of temporal variance of methylation between the two groups of mothers, amongst which the one with the largest effect belonged to the gene coding Uronyl 2-Sulfotransferase (*UST*). A previous study on gene expression analysis of peripheral blood in preterm delivering women compared those delivering at term reported a 2.05 fold higher expression of *UST* [38]. Our results probably indicate that the longitudinal variability of promoter DNA methylation of this gene might be the underlying cause of this observation.

Placenta mediates rapid physiological changes in pregnancy. During later part of pregnancy, certain genes which are expressed in placenta during mid-gestation, remain highly expressed until term. One such gene is *BMP5* (Bone morphogenetic protein 5), which showed higher temporal variance in preterm delivering women. It has been observed earlier that *BMP5* gene expression in placenta peaks in the second trimester (58.89 fold) compared to the first trimester, followed by a sharp decline at term in healthy term deliveries [39]. Studies have associated non-significant differential expression of *BMP5* gene in pregnancy related complications like preeclampsia, gestational diabetes, including small for gestational age as well as large for gestational age in the mid-gestation placenta. *BMP5* is implicated in bone and cartilage related disorders such as rheumatoid arthritis and osteoarthritis, also known to contribute to fetal development and growth. Probably, the variability in DNA methylation pattern of BMP5 detected in our study might serve to be a PTB specific signature in maternal blood. However, strictly to maternal peripheral blood specific signature for PTB.

A substantial proportion of the genes (20.69%, corresponding to 12 genes) which were found to exhibit temporal variance in DNAm in preterm birth delivering mothers in this study to the Platelet-derived growth factor (PDGF) signaling pathway. PDGF is a major mitogen for fibroblasts, smooth muscle cells, and other cells [40]. Both PDGF and PDGF receptor (PDGFR) expression patterns are spatio-temporally regulated during development and physiological processes [41,42]. Mesenchymal expression of PDGFRs is low in vivo. Several factors induce PDGFR expression, including TGF-β, estrogen (probably linked to hypertrophic smooth muscle responses in the pregnant uterus), interleukin-1α (IL-1α), basic fibroblast growth factor-2 (FGF-2), tumor necrosis factor-α, and lipopolysaccharide [43]. Furthermore, increased expression of PDGF-A is observed in human uterine smooth muscle cells during the physiological hypertrophy of pregnancy [44]. Platelet turnover is usually within normal range during pregnancy, except during the third trimester when gestational thrombocytopenia sets in, which is associated with higher rate of cesarean delivery compared to women who do not have this condition[45,46]. Increased platelet aggregation has been observed during pregnancy [47]. Platelet activation and simultaneous release of vasoactive substances occur especially during repair of the utero-placental unit during obstetric complications [48]. Variability in promoter DNA methylation of PDGF signaling genes could lead to variable expression which in turn might result in adverse outcomes like PTB by hindering downstream recruitment of vasoactive utero-placental repair.

Successful pregnancy is established by a series of physiological events including breakdown and remodelling of the extracellular matrix (ECM) in the endometrium. ECM proteoglycans play key roles in maintaining the connective tissue matrix and tensile strength of human fetal membranes and have been previously linked to pPROM (preterm premature rupture of membranes) [49]. ECM proteoglycans like decorin and biglycan act as biomarkers for increased risk of PPROM risk in maternal blood [49]. Excessive deposition of ECM in the connective tissue has also been associated with preeclampsia [50]. PPROM and preterm labor are discrete pathologic processes that can lead to spontaneous PTB. There is much overlap in the etiologic factors that predispose to these conditions [51], as well as exhibit much overlap in their clinical presentations. Alterations of DNA methylation leading of the ECM proteoglycans pathway genes leading to changes in their expression may therefore potentially explain their involvement in PTB outcome.

The primary component of ECM is collagen, which helps in endothelial cell proliferation, migration and differentiation, in order to form new vascular networks. Extensive collagen metabolism facilitates cervical remodelling. Cervical softening, shortening and dilation caused by early cervical remodelling can lead to spontaneous preterm birth(sPTB) [52]. The abundance of collagen in the fetal membranes decreases with gestational age [53,54]. Collagen degradation is initially brought by collagenase which is synthesized by cervical fibroblasts shortly before term delivery and activated during labor. Collagen degradation plays an essential role in prostaglandin (PG)-induced cervical ripening and the physiological maturation of the cervix in late pregnancy and at term [55,56]. Activity of collagen at the maternal fetal interface is therefore dynamic during the establishment of pregnancy. Interestingly, some of the genes which we identified belonged to the collagen degradation pathway. We hypothesize that temporal variance in the expression of these genes, as evidenced from the DNA methylation changes, might lead to preterm delivery.

Pregnancy is considered as a period susceptible to chronodisruption as pregnant women often undergo physiological, social and emotional changes, which may alter their usual lifestyle. Chronodisruption can occur either due to polymorphisms in the core clock genes [57], or pregnant women can experience environmental factors that force them to sleep or wake at times different than their biological clock. Furthermore, nutrition, physical activity, and stress can also contribute to chronodisruption[58,59]. Such chronodisruptive factors can increase the risk of adverse pregnancy outcomes like miscarriage, low birth weight, and PTB [60–62]. Chronodisruption may affect pregnancy outcomes such as PTB, however, the interaction of maternal, fetal, and placental circadian systems to regulate gestational length is yet unclear. A study in a Canadian cohort have shown that lower levels of transcript of the circadian clock gene Cryptochrome 2 (*CRY2*) in second trimester peripheral blood from mothers delivering preterm compared to term delivering mothers, might be associated with an increased risk of sPTB [63]. The same study also identified degradation of the ECM as one of the most significant pathways that could potentially lead to sPTB. Our study shows similar results with the DNAm signatures in the Indian cohort, thereby particularly establishing the importance and relevance of maternal component towards preterm delivery.

The importance of the impact of maternal circadian rhythm on pregnancy and fetal development is only now being recognized. It has been proposed that the development of fetal suprachiasmatic nuclei (SCN) and fetal organs are entrained by maternal circadian oscillators which results in temporal order during fetal growth. This subsequently allows for postnatal integration of an adult-like circadian clock in the fetal system[64]. Further investigations in-depth are needed to fully comprehend the effect of chronodisruption on risk of preterm birth and its effect on infant health.

To our knowledge this is the first study that focuses on the associations between temporal DNAm levels and PTB in specifically an Indian cohort. Although novel, our study has certain limitations. This is a discovery study and demands subsequent validation of the findings in an independent group of mothers. Additionally, the mechanistic follow-up of our findings need to be undertaken, which could help in providing deep understanding of the biological implications.

This study has several limitations that should be considered when interpreting the findings. First, the lack of pre-pregnancy DNAm levels, which would allow us to examine the role of the change in these levels during the course of pregnancy in PTB. Second, our sample size was relatively small, thus future well-powered studies are needed in order to perform validation analyses which could shed more light on the DNAm variability that exist in the preterm delivering mothers compared to term delivery. To our knowledge this is the first study that focuses on the associations between temporal DNAm levels and PTB in specifically an Indian cohort.

## Conclusion

We have tested our hypothesis that large variations in levels of genome-wide methylation correlate with preterm birth, using a novel method of analysis of longitudinal genome wide DNA methylation data in groups of individuals with contrasting pregnancy outcomes. Our study identified 1197 genes which exhibited significantly higher temporal variance in mothers delivering preterm as compared to those delivering at term. The results of our study, which is the first of its kind on maternal peripheral blood, has the potential to contribute towards improved understanding of PTB and might lead to development of a reduced invasive method of predicting risk of preterm delivery. We plan to validate our results in a larger set of mothers enrolled in GARBH-Ini, in order to to identify DNAm biomarkers which can be used for detection of PTB earlier during pregnancy.

## Data Availability

The DNA methylation data of this study and the source codes are openly available in Gene Expression Omnibus at https://www.ncbi.nlm.nih.gov/geo/query/acc.cgi?acc=GSE169338, under the study accession number GSE169338 and GitHub: https://github.com/Jagyashila/ptbmethylation respectively.

## Acknowledgements

Professor M. K. Bhan will always be remembered reverently for his critical scientific and technical feedback. The authors recognize the efforts of the personnel of the GARBH-Ini Cohort and laboratory support from the Core Technologies Research Initiative (CoTeRI). We would like to thank Sahana Ghosh of NIBMG for assistance on submission of the data in GEO. This work was funded by the Department of Biotechnology, Ministry of Science and Technology, Govt. of India (BT/PR9983/MED/97/194/2013) and for some components of the biorepository by the Grand Challenges India – All Children Thriving Program (supported by the Programme Management Unit), Biotechnology Industry Research Assistance Council (BIRAC/GCI/0114/03/14–ACT). PPM acknowledges support of his J. C. Bose Fellowship and National Science Chair. JD is supported by the Research Fellowship (NET) of the University Grants Commission (UGC), India. The authors are thankful to Mr. Sukhdev Sinha and Dr. Anamika Gambhir for their technical feedback and support.

