## Supplementary Information for "Pattern of variation in DNA methylation during pregnancy among mothers who delivered preterm in the GARBH-Ini cohort"

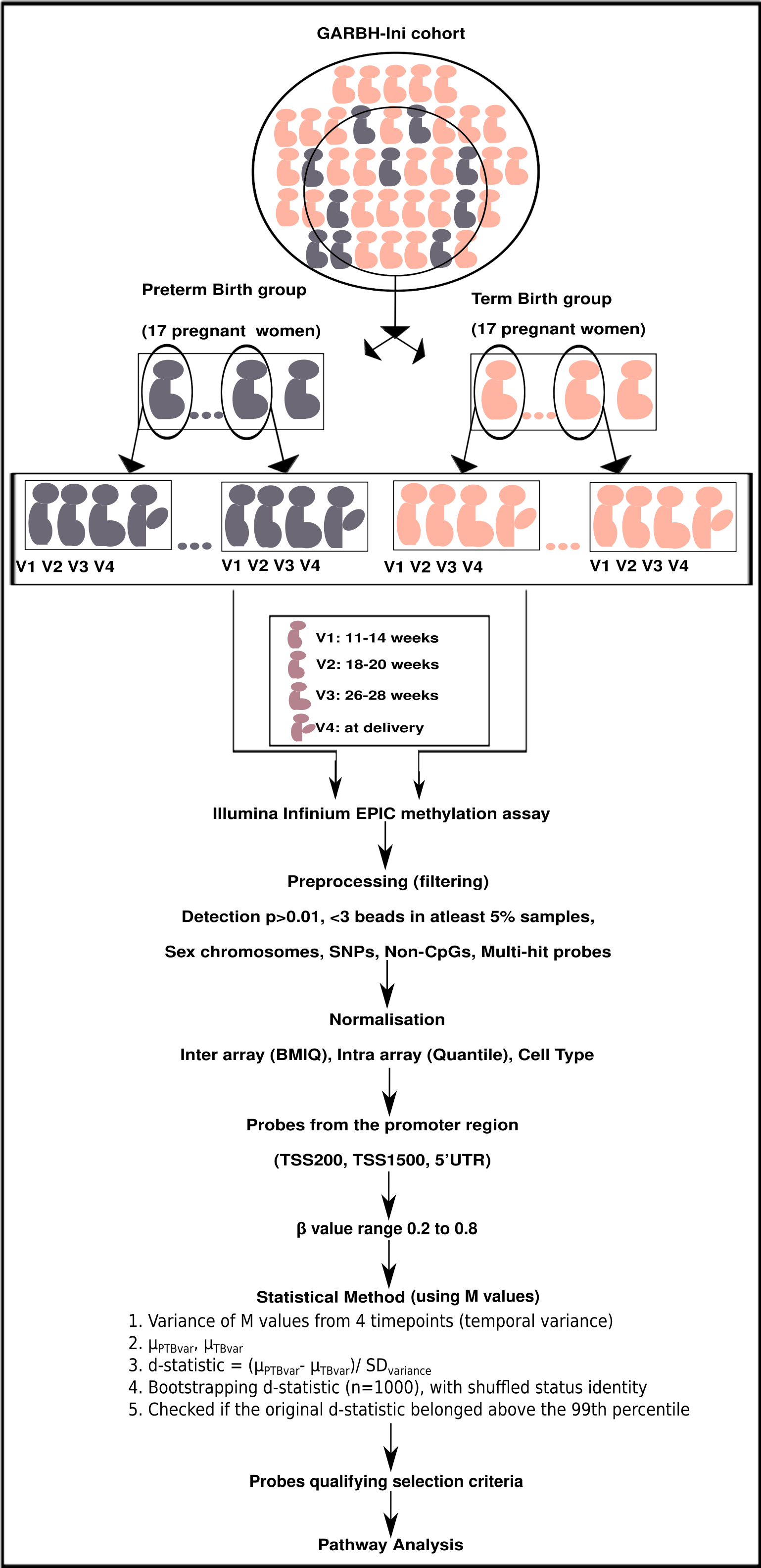

**Supplementary Fig 1:** Workflow for analysis of temporal DNA methylation data from term and preterm delivering women, from the GARBH-Ini cohort.

| **Criteria** | **No.of probes removed** | **No. of probes retained** |
| --- | --- | --- |
| **Preprocessing(red and green channel intensity >3000, detection P <0.01, <3 beads in atleast 5% samples)** | 3700 | 862218 |
| **Sex chromosomes** | 17844 | 844374 |
| **SNPs** | 81493 | 762881 |
| **Non-CpGs** | 2991 | 759890 |
| **Multi-Hit probes** | 49 | 759841 |
| **Probes from promoter regions (TSS200, TSS1500, 5’UTR)** | 540677 | 219164 |
| **M value range -2 to 2** | 193056 | **26108** |

**Supplementary Table 1:** Steps undertaken as Quality Control and probe selection criteria.

| **Participant ID** | **POG V1 (days)** | **POG V2 (days)** | **POG V3 (days)** | **POG V5 (days)** |
| --- | --- | --- | --- | --- |
| 0345 | 59 | 131 | 186 | 276 |
| 0735 | 90 | 135 | 184 | 228 |
| 0733 | 84 | 133 | 183 | 291 |
| 0952 | 91 | 130 | 183 | 241 |
| 1131 | 95 | 131 | 183 | 277 |
| 1085 | 50 | 130 | 183 | 235 |
| 1378 | 58 | 130 | 183 | 281 |
| 1423 | 48 | 130 | 183 | 244 |
| 1031 | 42 | 131 | 185 | 279 |
| 1537 | 75 | 131 | 185 | 190 |
| 3138 | 66 | 130 | 184 | 268 |
| 3248 | 57 | 130 | 186 | 231 |
| 3501 | 85 | 132 | 192 | 288 |
| 3640 | 77 | 130 | 183 | 232 |
| 4006 | 94 | 130 | 184 | 276 |
| 4293 | 85 | 133 | 186 | 220 |
| 0029 | 48 | 130 | 187 | 268 |
| 0107 | 88 | 132 | 187 | 247 |
| 0203 | 70 | 130 | 185 | 277 |
| 0285 | 90 | 137 | 184 | 244 |
| 0375 | 82 | 128 | 194 | 259 |
| 0464 | 50 | 132 | 183 | 206 |
| 0828 | 88 | 133 | 183 | 276 |
| 0978 | 90 | 130 | 183 | 248 |
| 1213 | 43 | 136 | 183 | 267 |
| 1441 | 61 | 130 | 184 | 244 |
| 0682 | 86 | 132 | 188 | 263 |
| 0876 | 73 | 130 | 184 | 190 |
| 0251 | 60 | 136 | 184 | 266 |
| 0328 | 83 | 131 | 184 | 251 |
| 1563 | 67 | 132 | 184 | 279 |
| 1656 | 58 | 134 | 187 | 250 |
| 1769 | 94 | 145 | 183 | 273 |
| 1845 | 98 | 130 | 183 | 244 |

**Supplementary Table 2:** Time of collection of peripheral blood samples from study participants, estimated in days from the time of conception using ultrasonography.

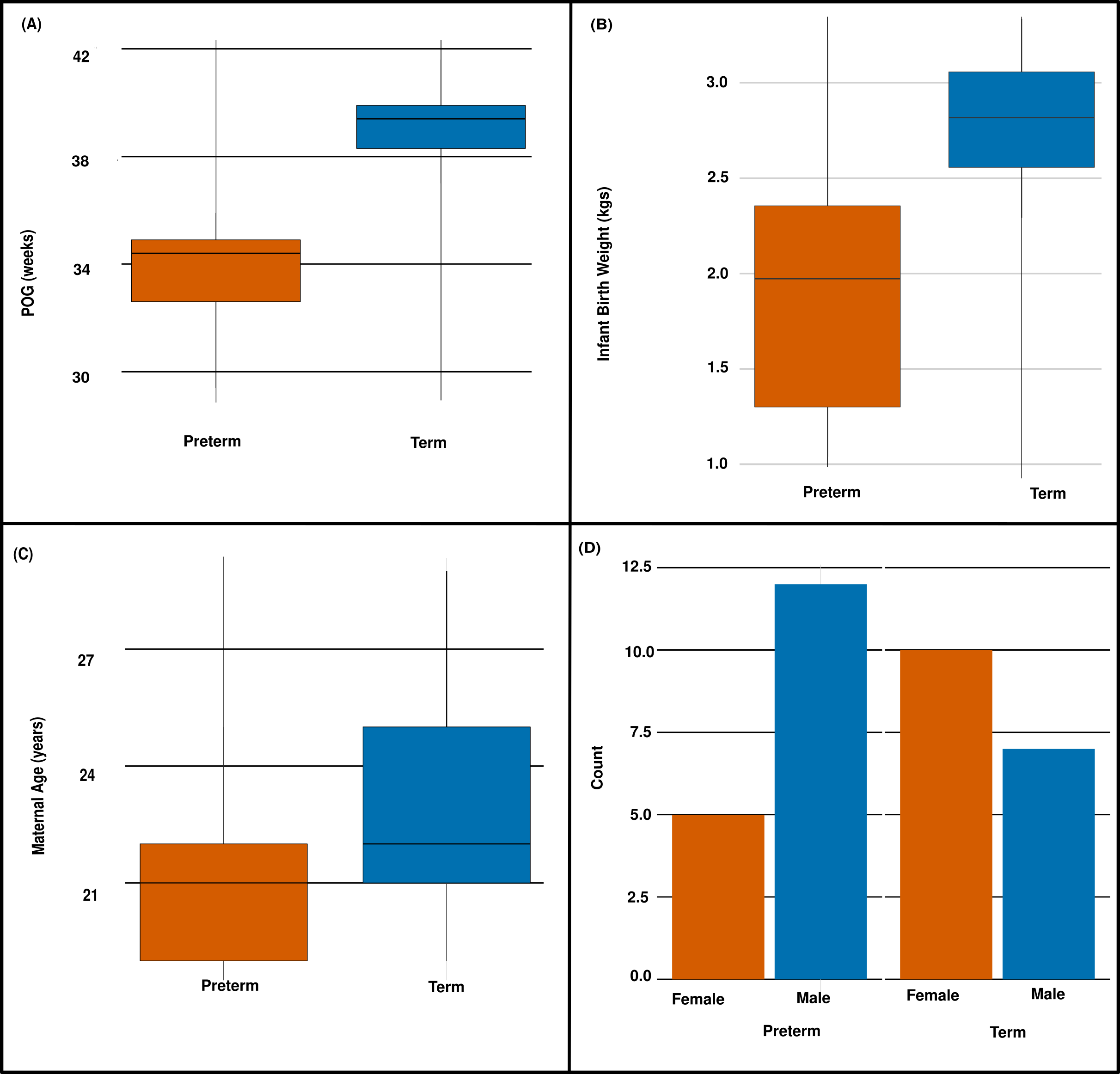

**Supplementary Fig 2:** Boxplots showing (A) period of gestation (POG) in weeks between preterm and term delivering mothers; (B) infant birth weight measured at delivery, between preterm and term delivery; (C) age of preterm and term delivering women; and, (D) Bar plots showing number of male and female infant born preterm and term.

**Supplementary table 3:** List of probes (1296) with standardized difference in mean temporal variance between the preterm and term delivering mothers above 99th percentile.

**Supplementary table 4:** List of probes (1296) that belonged to the highest 5% of all probes with significant mean difference in temporal variance of methylation between mothers who delivered at term and preterm

| **Sl No.** | **Gene Name** | **CpG ID** | **d-statistic** |
| --- | --- | --- | --- |
| 1 | UST | cg03366867 | 1.77 |
| 2 | OSBPL3 | cg00891520 | 1.63 |
| 3 | CGNL1 | cg26185468 | 1.58 |
| 4 | TBC1D24 | cg08190844 | 1.58 |
| 5 | NFYC | cg26783428 | 1.52 |
| 6 | ZNF165 | cg06532659 | 1.5 |
| 7 | BMP5 | cg24926954 | 1.47 |
| 8 | CKM | cg20444256 | 1.47 |
| 9 | C5orf47 | cg21756326 | 1.47 |
| 10 | TNRC6B | cg21034183 | 1.45 |
| 11 | CLK3 | cg05480045 | 1.44 |
| 12 | ASNSD1 | cg16484820 | 1.41 |
| 13 | DNAJC3 | cg08599294 | 1.4 |
| 14 | IBTK | cg03142554 | 1.39 |
| 15 | VDR | cg02547054 | 1.39 |
| 16 | OR6C4 | cg10164401 | 1.37 |
| 17 | TNFRSF9 | cg18964839 | 1.37 |
| 18 | KRTAP5-3 | cg23831386 | 1.36 |
| 19 | SCOC | cg04761746 | 1.36 |
| 20 | MIR31HG | cg00632587 | 1.35 |
| 21 | LINC00222 | cg11817818 | 1.34 |
| 22 | BRPF3 | cg13719287 | 1.34 |
| 23 | C1QTNF3 | cg23051392 | 1.33 |
| 24 | ALLC | cg19825600 | 1.32 |
| 25 | AKR1C4 | cg11894854 | 1.32 |
| 26 | TMC5 | cg24144243 | 1.31 |
| 27 | DDIAS | cg14597502 | 1.3 |
| 28 | PTP4A2 | cg08757017 | 1.3 |
| 29 | MIR320C1 | cg02645550 | 1.28 |
| 30 | GPRC5B | cg09141413 | 1.28 |
| 31 | C5orf13 | cg22562498 | 1.28 |
| 32 | NCOA2 | cg25986804 | 1.27 |
| 33 | PTPRQ | cg15535380 | 1.27 |
| 34 | ORAI1 | cg02827278 | 1.27 |
| 35 | PRKG2 | cg11283514 | 1.27 |
| 36 | DOPEY1 | cg14756497 | 1.26 |
| 37 | ARHGEF10 | cg06463230 | 1.26 |
| 38 | XPOT | cg12609893 | 1.26 |
| 39 | NR2C2 | cg09438522 | 1.25 |
| 40 | FAM110A | cg00622595 | 1.24 |
| 41 | TAGLN | cg16524733 | 1.23 |
| 42 | SEMA6D | cg20611207 | 1.23 |
| 43 | PIGY | cg18521726 | 1.23 |
| 44 | JADE1 | cg01986502 | 1.23 |
| 45 | TNNT2 | cg02589576 | 1.23 |
| 46 | TREML1 | cg15153887 | 1.22 |
| 47 | TNK2 | cg23548201 | 1.22 |
| 48 | SRPK2 | cg01150173 | 1.22 |
| 49 | LDLRAD3 | cg17684006 | 1.22 |
| 50 | KRTAP19-4 | cg06515562 | 1.22 |
| 51 | CACNG4 | cg16116425 | 1.22 |
| 52 | ZNF532 | cg26172016 | 1.21 |
| 53 | HTR6 | cg04607131 | 1.21 |
| 54 | TMC5 | cg21800306 | 1.2 |
| 55 | ADGRG1 | cg10696060 | 1.2 |
| 56 | SEMA4D | cg00437441 | 1.2 |
| 57 | AIG1 | cg26552913 | 1.2 |
| 58 | PARVG | cg14942952 | 1.2 |
| 59 | OR2T4 | cg10464163 | 1.2 |
| 60 | DLL3 | cg20077879 | 1.19 |
| 61 | LOC101929680 | cg15564949 | 1.19 |
| 62 | PAFAH1B3 | cg21810808 | 1.19 |
| 63 | TRAK1 | cg08508763 | 1.19 |
| 64 | A2ML1 | cg23546356 | 1.19 |
| 65 | RBPMS | cg26122129 | 1.19 |
| 66 | ANKRD53 | cg16612112 | 1.18 |
| 67 | ZSCAN10 | cg22520445 | 1.18 |
| 68 | TSPO | cg24710387 | 1.18 |
| 69 | TREM2 | cg02828883 | 1.18 |
| 70 | RFESD | cg22136351 | 1.17 |
| 71 | RAB32 | cg01892997 | 1.17 |
| 72 | ADAM33 | cg00548989 | 1.17 |
| 73 | GPR146 | cg24920909 | 1.17 |
| 74 | OR9Q1 | cg02666556 | 1.17 |
| 75 | NDUFC1 | cg04006143 | 1.17 |
| 76 | COQ6 | cg16524108 | 1.17 |
| 77 | FILIP1L | cg23221889 | 1.17 |
| 78 | TLE2 | cg11672493 | 1.17 |
| 79 | TEAD3 | cg20595752 | 1.16 |
| 80 | RORC | cg10269078 | 1.16 |
| 81 | ZNF790 | cg04803153 | 1.16 |
| 82 | ZNF438 | cg23260901 | 1.16 |
| 83 | TPST2 | cg20682816 | 1.16 |
| 84 | LCE2B | cg25098401 | 1.16 |
| 85 | LPPR1 | cg14253327 | 1.16 |
| 86 | TMEM220 | cg11750736 | 1.16 |
| 87 | RUNX1T1 | cg19007731 | 1.16 |
| 88 | SPSB1 | cg15039460 | 1.15 |
| 89 | KLK11 | cg09702010 | 1.15 |
| 90 | CSF2 | cg13103915 | 1.15 |
| 91 | SLC26A8 | cg00770599 | 1.15 |
| 92 | LINC00397 | cg06681689 | 1.15 |
| 93 | TBK1 | cg16604658 | 1.15 |
| 94 | ARHGAP22 | cg21586764 | 1.14 |
| 95 | OVCH2 | cg25499650 | 1.14 |
| 96 | R3HDM1 | cg11852007 | 1.14 |
| 97 | MDC1 | cg10925640 | 1.14 |
| 98 | ABHD10 | cg10672681 | 1.14 |
| 99 | LCN6 | cg13544075 | 1.13 |
| 100 | IGDCC4 | cg07617246 | 1.13 |
| 101 | CD27 | cg22395761 | 1.13 |
| 102 | CLPS | cg23264899 | 1.13 |
| 103 | CREB5 | cg21581567 | 1.13 |
| 104 | DYRK4 | cg10559778 | 1.13 |
| 105 | MMAA | cg06788644 | 1.13 |
| 106 | LOC101927844 | cg07642404 | 1.13 |
| 107 | BPIFA2 | cg05537333 | 1.13 |
| 108 | RPL24 | cg00653460 | 1.12 |
| 109 | HSPA1L | cg17145983 | 1.12 |
| 110 | DTNA | cg11890793 | 1.12 |
| 111 | MGC57346-CRHR1 | cg02833950 | 1.12 |
| 112 | GAS7 | cg14257839 | 1.12 |
| 113 | AP2B1 | cg08865496 | 1.11 |
| 114 | LOC285768 | cg04186487 | 1.11 |
| 115 | SGMS2 | cg14616533 | 1.11 |
| 116 | TLE2 | cg24004577 | 1.11 |
| 117 | BCHE | cg21995126 | 1.11 |
| 118 | UBALD2 | cg11278212 | 1.11 |
| 119 | DPEP1 | cg18555348 | 1.11 |
| 120 | HSPB7 | cg03260781 | 1.11 |
| 121 | PCDHGB6 | cg13254082 | 1.11 |
| 122 | NARS | cg08648241 | 1.11 |
| 123 | C12orf59 | cg03077730 | 1.11 |
| 124 | CCSER1 | cg22722548 | 1.11 |
| 125 | CNFN | cg17107617 | 1.1 |
| 126 | PSMB2 | cg24109894 | 1.1 |
| 127 | CIZ1 | cg14217552 | 1.1 |
| 128 | RPL36 | cg23630452 | 1.1 |
| 129 | HLA-J | cg21723245 | 1.1 |
| 130 | SNRPN | cg19803984 | 1.1 |
| 131 | MXI1 | cg15054954 | 1.1 |
| 132 | PCDH1 | cg21840108 | 1.1 |
| 133 | C2orf63 | cg25477606 | 1.1 |
| 134 | PHGDH | cg05958127 | 1.1 |
| 135 | KRT28 | cg16048803 | 1.1 |
| 136 | IFT81 | cg03698892 | 1.1 |
| 137 | PCGF3 | cg08449090 | 1.1 |
| 138 | LRP4 | cg23062006 | 1.09 |
| 139 | IFNL4 | cg26680763 | 1.09 |
| 140 | PPFIA4 | cg13884889 | 1.09 |
| 141 | FBLIM1 | cg25719573 | 1.09 |
| 142 | FLJ25758 | cg01993907 | 1.09 |
| 143 | ELOVL3 | cg00431050 | 1.09 |
| 144 | CDIPT | cg22191015 | 1.09 |
| 145 | TMCC1 | cg19405504 | 1.09 |
| 146 | KANK1 | cg15828854 | 1.09 |
| 147 | MANBA | cg03749037 | 1.09 |
| 148 | FRS2 | cg15736548 | 1.09 |
| 149 | KLF1 | cg22894983 | 1.09 |
| 150 | GSTO1 | cg08262530 | 1.09 |
| 151 | OXT | cg13285174 | 1.09 |
| 152 | EYA2 | cg07899984 | 1.08 |
| 153 | CTSB | cg16687420 | 1.08 |
| 154 | KIAA1609 | cg09259081 | 1.08 |
| 155 | ZNF784 | cg24053609 | 1.08 |
| 156 | SYCP1 | cg07832006 | 1.08 |
| 157 | LGALS8 | cg09334514 | 1.08 |
| 158 | XRCC3 | cg19958417 | 1.08 |
| 159 | TTLL6 | cg06169270 | 1.08 |
| 160 | MEI1 | cg23592953 | 1.08 |
| 161 | RGR | cg00415971 | 1.08 |
| 162 | ZMAT4 | cg17635352 | 1.08 |
| 163 | HOXA3 | cg23403004 | 1.08 |
| 164 | CYTIP | cg03915055 | 1.08 |
| 165 | RTKN | cg14481208 | 1.08 |
| 166 | GID4 | cg03727280 | 1.07 |
| 167 | STON1-GTF2A1L | cg07289618 | 1.07 |
| 168 | LOC101929771 | cg13703134 | 1.07 |
| 169 | MAP2K3 | cg13139135 | 1.07 |
| 170 | SLC39A7 | cg17567838 | 1.07 |
| 171 | TMEM8B | cg15818306 | 1.07 |
| 172 | C16orf67 | cg09713758 | 1.07 |
| 173 | KLHL3 | cg18589597 | 1.07 |
| 174 | TGFBR3 | cg00342358 | 1.07 |
| 175 | MECOM | cg10358466 | 1.06 |
| 176 | SLA2 | cg23779478 | 1.06 |
| 177 | SLC39A10 | cg26970043 | 1.06 |
| 178 | SEMA7A | cg20110364 | 1.06 |
| 179 | PLAT | cg12765042 | 1.06 |
| 180 | TUBB | cg27505349 | 1.06 |
| 181 | FXYD4 | cg24294544 | 1.06 |
| 182 | LOC101929563 | cg22609575 | 1.06 |
| 183 | MYLK | cg10935889 | 1.06 |
| 184 | PIANP | cg03052567 | 1.06 |
| 185 | CTNNA2 | cg13711653 | 1.06 |
| 186 | CHRM2 | cg07172149 | 1.06 |
| 187 | AADAT | cg09371608 | 1.06 |
| 188 | C19orf41 | cg03095288 | 1.06 |
| 189 | C1QA | cg00291765 | 1.06 |
| 190 | KIAA1529 | cg13713218 | 1.06 |
| 191 | MUC6 | cg07407723 | 1.06 |
| 192 | FLJ45079 | cg04837616 | 1.06 |
| 193 | DPP9 | cg15404665 | 1.05 |
| 194 | MIR1470 | cg14892570 | 1.05 |
| 195 | FGR | cg13595076 | 1.05 |
| 196 | KANSL1 | cg17003784 | 1.05 |
| 197 | NSUN4 | cg24483800 | 1.05 |
| 198 | LARGE | cg01741606 | 1.05 |
| 199 | RBFOX1 | cg14874133 | 1.05 |
| 200 | GPR148 | cg15196820 | 1.05 |
| 201 | FANCC | cg09394661 | 1.05 |
| 202 | SYT3 | cg06732870 | 1.05 |
| 203 | RGS3 | cg13066954 | 1.05 |
| 204 | RNFT2 | cg03109701 | 1.05 |
| 205 | GPER | cg06449934 | 1.05 |
| 206 | FOXI2 | cg25043050 | 1.05 |
| 207 | EXOC3 | cg05387362 | 1.05 |
| 208 | CNTN1 | cg26359940 | 1.05 |
| 209 | FERMT1 | cg07718903 | 1.05 |
| 210 | S100B | cg07152925 | 1.04 |
| 211 | CARD11 | cg19014792 | 1.04 |
| 212 | SLC38A11 | cg12301695 | 1.04 |
| 213 | ZNF841 | cg05974498 | 1.04 |
| 214 | NEDD9 | cg14634313 | 1.04 |
| 215 | NPPB | cg17641943 | 1.04 |
| 216 | MYH8 | cg10798382 | 1.04 |
| 217 | MMP19 | cg16936370 | 1.04 |
| 218 | CCDC149 | cg09490880 | 1.04 |
| 219 | COL12A1 | cg03503642 | 1.04 |
| 220 | SCN1A | cg05281304 | 1.04 |
| 221 | PRKCD | cg08170911 | 1.04 |
| 222 | SLC45A3 | cg15826544 | 1.04 |
| 223 | RSPH3 | cg26011786 | 1.04 |
| 224 | MGC57346-CRHR1 | cg17509718 | 1.04 |
| 225 | C4orf23 | cg21641512 | 1.04 |
| 226 | DAB1 | cg22076476 | 1.04 |
| 227 | AGPAT3 | cg23504415 | 1.04 |
| 228 | GFY | cg06744472 | 1.04 |
| 229 | BTG4 | cg09148270 | 1.04 |
| 230 | ELOVL5 | cg09387914 | 1.04 |
| 231 | C10orf128 | cg00142482 | 1.04 |
| 232 | BCL6 | cg15232708 | 1.04 |
| 233 | TEAD1 | cg08419476 | 1.03 |
| 234 | CHST4 | cg20077275 | 1.03 |
| 235 | BCO2 | cg08355483 | 1.03 |
| 236 | RANBP1 | cg18631743 | 1.03 |
| 237 | SLC38A11 | cg16376155 | 1.03 |
| 238 | CHL1-AS1 | cg11091882 | 1.03 |
| 239 | TECRL | cg21482647 | 1.03 |
| 240 | OR2AE1 | cg20129838 | 1.03 |
| 241 | MVB12A | cg25211050 | 1.03 |
| 242 | NKIRAS2 | cg22361181 | 1.03 |
| 243 | TLR9 | cg18452449 | 1.03 |
| 244 | FAM123A | cg00313750 | 1.03 |
| 245 | LDLRAD4 | cg26745882 | 1.03 |
| 246 | ANXA6 | cg10610249 | 1.03 |
| 247 | ATP6V1C1 | cg14341424 | 1.03 |
| 248 | BBOX1 | cg08185591 | 1.02 |
| 249 | SLC4A5 | cg20336730 | 1.02 |
| 250 | TRABD2B | cg06701732 | 1.02 |
| 251 | RASGRP4 | cg18007970 | 1.02 |
| 252 | PNOC | cg10416846 | 1.02 |
| 253 | TBC1D23 | cg16920250 | 1.02 |
| 254 | NEDD9 | cg01700428 | 1.02 |
| 255 | FAM84A | cg22044566 | 1.02 |
| 256 | LTB4R | cg09274091 | 1.02 |
| 257 | OIT3 | cg13817271 | 1.02 |
| 258 | MAPK12 | cg04956306 | 1.02 |
| 259 | SLC25A1 | cg22766963 | 1.02 |
| 260 | C11orf94 | cg04310460 | 1.02 |
| 261 | LYRM1 | cg06126593 | 1.02 |
| 262 | NCR1 | cg11356156 | 1.02 |
| 263 | ASPA | cg18769729 | 1.02 |
| 264 | C2 | cg27297326 | 1.02 |
| 265 | GOLSYN | cg13456086 | 1.02 |
| 266 | LY86 | cg07910479 | 1.02 |
| 267 | ADAM33 | cg22333888 | 1.02 |
| 268 | C7orf57 | cg20674327 | 1.02 |
| 269 | MOSPD3 | cg21063500 | 1.02 |
| 270 | C6orf35 | cg22491629 | 1.02 |
| 271 | ZBTB16 | cg09360036 | 1.02 |
| 272 | BMP8B | cg06616202 | 1.02 |
| 273 | FSCB | cg01119259 | 1.02 |
| 274 | ST5 | cg25831522 | 1.02 |
| 275 | SORBS2 | cg04348265 | 1.01 |
| 276 | TIAF1 | cg25743584 | 1.01 |
| 277 | PLEKHG5 | cg04434328 | 1.01 |
| 278 | TMCO7 | cg01650965 | 1.01 |
| 279 | LOC285033 | cg04232816 | 1.01 |
| 280 | DICER1 | cg24293739 | 1.01 |
| 281 | CPNE6 | cg03523676 | 1.01 |
| 282 | SMARCA2 | cg15067568 | 1.01 |
| 283 | CLDN15 | cg08636573 | 1.01 |
| 284 | NUDT2 | cg16532903 | 1.01 |
| 285 | DMKN | cg17112426 | 1.01 |
| 286 | FAM111B | cg21675569 | 1.01 |
| 287 | GIPR | cg14661225 | 1.01 |
| 288 | EPHA3 | cg18055394 | 1.01 |
| 289 | TTYH2 | cg23327200 | 1 |
| 290 | TCF7 | cg08063911 | 1 |
| 291 | RB1CC1 | cg02211317 | 1 |
| 292 | FSHR | cg19927457 | 1 |
| 293 | INCA1 | cg09578319 | 1 |
| 294 | MIR548C | cg16098340 | 1 |
| 295 | SNRPB2 | cg24899750 | 1 |
| 296 | APOC3 | cg20957428 | 1 |
| 297 | MICAL3 | cg00040575 | 1 |
| 298 | ANKRD18B | cg00054209 | 1 |
| 299 | FAM104A | cg25559913 | 1 |
| 300 | DGKQ | cg23634348 | 1 |
| 301 | SH3BP4 | cg09943385 | 1 |
| 302 | SUPT3H | cg01946401 | 1 |
| 303 | C14orf135 | cg08082389 | 1 |
| 304 | APIP | cg21916100 | 1 |
| 305 | RAI1 | cg15928524 | 1 |
| 306 | PHKG1 | cg21940640 | 1 |
| 307 | ZNF540 | cg20700792 | 1 |
| 308 | PHF1 | cg03157610 | 1 |
| 309 | PEF1 | cg20910528 | 1 |
| 310 | LIMCH1 | cg02578871 | 0.99 |
| 311 | C6orf146 | cg11744524 | 0.99 |
| 312 | ZNF746 | cg02401978 | 0.99 |
| 313 | DKFZp779M0652 | cg05923914 | 0.99 |
| 314 | LSM4 | cg06004060 | 0.99 |
| 315 | GATAD2A | cg12488919 | 0.99 |
| 316 | SCNN1A | cg09680149 | 0.99 |
| 317 | C3orf1 | cg00261569 | 0.99 |
| 318 | ANO8 | cg03665970 | 0.99 |
| 319 | CRABP1 | cg02260363 | 0.99 |
| 320 | YWHAG | cg23214628 | 0.99 |
| 321 | ALDH1L2 | cg20589117 | 0.99 |
| 322 | ACER3 | cg02208432 | 0.99 |
| 323 | PCDHA11 | cg19239201 | 0.99 |
| 324 | PPP1CA | cg01789162 | 0.99 |
| 325 | SNORD116-24 | cg09299132 | 0.99 |
| 326 | CPLX2 | cg15301006 | 0.99 |
| 327 | DAPL1 | cg02251393 | 0.99 |
| 328 | NDUFS2 | cg07580762 | 0.99 |
| 329 | OPALIN | cg26268085 | 0.99 |
| 330 | MFAP2 | cg14657514 | 0.99 |
| 331 | CCNY | cg23460832 | 0.99 |
| 332 | SLC16A3 | cg10208942 | 0.99 |
| 333 | GRB2 | cg17632777 | 0.99 |
| 334 | S100Z | cg05199418 | 0.99 |
| 335 | UNKL | cg14850784 | 0.99 |
| 336 | DFNA5 | cg20764575 | 0.99 |
| 337 | UBB | cg00883384 | 0.99 |
| 338 | GABRG2 | cg04550524 | 0.99 |
| 339 | RPH3AL | cg25255204 | 0.99 |
| 340 | C6orf227 | cg00536532 | 0.99 |
| 341 | SERTAD4 | cg13884907 | 0.99 |
| 342 | MTHFD2P1 | cg21254120 | 0.99 |
| 343 | HAMP | cg02131995 | 0.99 |
| 344 | PGAP1 | cg00734053 | 0.99 |
| 345 | IRF8 | cg18651546 | 0.99 |
| 346 | GP1BA | cg10763547 | 0.98 |
| 347 | PCSK7 | cg00975249 | 0.98 |
| 348 | KCNAB2 | cg11740026 | 0.98 |
| 349 | PARP11 | cg03755508 | 0.98 |
| 350 | C9orf66 | cg04498816 | 0.98 |
| 351 | TLR5 | cg15448366 | 0.98 |
| 352 | STAT3 | cg06270615 | 0.98 |
| 353 | TUBB | cg08680415 | 0.98 |
| 354 | PSMF1 | cg13791092 | 0.98 |
| 355 | OR5B2 | cg22015585 | 0.98 |
| 356 | LPIN2 | cg14443640 | 0.98 |
| 357 | DES | cg13227481 | 0.98 |
| 358 | MAP3K14 | cg00146864 | 0.98 |
| 359 | ZBTB20 | cg03779374 | 0.98 |
| 360 | SLC24A6 | cg00302340 | 0.98 |
| 361 | EMID1 | cg02152034 | 0.98 |
| 362 | LOC441897 | cg16966139 | 0.98 |
| 363 | LAMP1 | cg20882571 | 0.98 |
| 364 | EDAR | cg01124420 | 0.98 |
| 365 | ST3GAL6 | cg13377633 | 0.98 |
| 366 | PCDHGA3 | cg24828065 | 0.97 |
| 367 | C16orf73 | cg27105890 | 0.97 |
| 368 | CECR2 | cg16845108 | 0.97 |
| 369 | KCTD4 | cg08872252 | 0.97 |
| 370 | TRAK2 | cg15971559 | 0.97 |
| 371 | NMUR1 | cg03164561 | 0.97 |
| 372 | ZNF597 | cg09596866 | 0.97 |
| 373 | REEP5 | cg10291279 | 0.97 |
| 374 | PIP5KL1 | cg04959285 | 0.97 |
| 375 | CCK | cg21681168 | 0.97 |
| 376 | EDAR | cg11306744 | 0.97 |
| 377 | MOBKL2C | cg03345668 | 0.97 |
| 378 | SOCS7 | cg26283343 | 0.97 |
| 379 | FAM228B | cg20322447 | 0.97 |
| 380 | MIR7-3HG | cg00329803 | 0.97 |
| 381 | LOC102723330 | cg23691588 | 0.97 |
| 382 | RPL23AP64 | cg01675512 | 0.97 |
| 383 | PKIG | cg00622242 | 0.97 |
| 384 | WDR38 | cg10392895 | 0.97 |
| 385 | TAGAP | cg16180082 | 0.97 |
| 386 | C22orf25 | cg00390484 | 0.97 |
| 387 | DAPK1 | cg02058168 | 0.97 |
| 388 | CRISPLD2 | cg03296797 | 0.97 |
| 389 | LPCAT1 | cg20233130 | 0.97 |
| 390 | FRMD1 | cg01951188 | 0.97 |
| 391 | GOLGA4 | cg04961265 | 0.97 |
| 392 | NPHS1 | cg14327228 | 0.97 |
| 393 | ZNF165 | cg00185909 | 0.97 |
| 394 | PPP2R2B | cg01987925 | 0.97 |
| 395 | ENO1 | cg20536660 | 0.97 |
| 396 | PRKACB | cg07474678 | 0.97 |
| 397 | ROS1 | cg24267752 | 0.97 |
| 398 | BIRC8 | cg19995259 | 0.96 |
| 399 | MMP23A | cg08231710 | 0.96 |
| 400 | TCTE1 | cg09347824 | 0.96 |
| 401 | AVP | cg16536918 | 0.96 |
| 402 | ZSCAN22 | cg18393172 | 0.96 |
| 403 | SDSL | cg16303241 | 0.96 |
| 404 | F13A1 | cg21594557 | 0.96 |
| 405 | BMPR1B | cg09771641 | 0.96 |
| 406 | EPS8L1 | cg05738030 | 0.96 |
| 407 | SLC8A1 | cg03604075 | 0.96 |
| 408 | NME4 | cg16018533 | 0.96 |
| 409 | MYO7B | cg22529396 | 0.96 |
| 410 | IMP3 | cg26746664 | 0.96 |
| 411 | GMPR | cg06476681 | 0.96 |
| 412 | CCDC114 | cg02744588 | 0.96 |
| 413 | C5AR2 | cg22422911 | 0.96 |
| 414 | FRYL | cg10109327 | 0.96 |
| 415 | PCDHB2 | cg11182018 | 0.96 |
| 416 | RACGAP1 | cg05088463 | 0.96 |
| 417 | APBA2 | cg03115989 | 0.96 |
| 418 | CEMIP | cg15857288 | 0.96 |
| 419 | GBE1 | cg11625933 | 0.96 |
| 420 | CCDC69 | cg17753843 | 0.96 |
| 421 | OR5T1 | cg01464104 | 0.96 |
| 422 | UPP1 | cg10213875 | 0.96 |
| 423 | LOC101929413 | cg15075483 | 0.96 |
| 424 | MRPL23 | cg09884480 | 0.96 |
| 425 | RAB7A | cg13404532 | 0.96 |
| 426 | TMEM105 | cg23514211 | 0.96 |
| 427 | SLC2A3 | cg02511315 | 0.95 |
| 428 | CEACAM21 | cg04946843 | 0.95 |
| 429 | CFAP44 | cg20032280 | 0.95 |
| 430 | SYNGR3 | cg26470314 | 0.95 |
| 431 | C3orf14 | cg15423991 | 0.95 |
| 432 | PTX4 | cg25921756 | 0.95 |
| 433 | CDH22 | cg07501939 | 0.95 |
| 434 | CCNH | cg27584762 | 0.95 |
| 435 | LCE2A | cg08166767 | 0.95 |
| 436 | KIAA0319L | cg16015113 | 0.95 |
| 437 | BAIAP3 | cg12708340 | 0.95 |
| 438 | GON4L | cg00504075 | 0.95 |
| 439 | TNXB | cg03543593 | 0.95 |
| 440 | GRIK4 | cg27438880 | 0.95 |
| 441 | TEAD4 | cg24713780 | 0.95 |
| 442 | UBAP1 | cg21231853 | 0.95 |
| 443 | SEMA4B | cg20246851 | 0.95 |
| 444 | SLC44A3 | cg20817062 | 0.95 |
| 445 | ZBTB32 | cg08539991 | 0.95 |
| 446 | RTDR1 | cg14458815 | 0.95 |
| 447 | VAT1L | cg15604403 | 0.95 |
| 448 | MYO1C | cg12741184 | 0.95 |
| 449 | HPCAL4 | cg00974095 | 0.95 |
| 450 | ZACN | cg04568564 | 0.95 |
| 451 | GPX4 | cg10732871 | 0.95 |
| 452 | NCAPD2 | cg10383428 | 0.95 |
| 453 | ATXN1 | cg26393261 | 0.95 |
| 454 | SIAH1 | cg08088511 | 0.95 |
| 455 | MAL2 | cg14221550 | 0.95 |
| 456 | ZBTB12 | cg24508713 | 0.94 |
| 457 | DMKN | cg00302312 | 0.94 |
| 458 | PPP1R13L | cg18766873 | 0.94 |
| 459 | NOL4 | cg10041241 | 0.94 |
| 460 | LPHN2 | cg25974355 | 0.94 |
| 461 | HBZ | cg01839639 | 0.94 |
| 462 | POU2F1 | cg05790989 | 0.94 |
| 463 | GAPDHS | cg15729137 | 0.94 |
| 464 | C1orf65 | cg26603179 | 0.94 |
| 465 | LOC100130015 | cg02434554 | 0.94 |
| 466 | HOXC11 | cg03762366 | 0.94 |
| 467 | LMTK3 | cg01354479 | 0.94 |
| 468 | LSP1 | cg08687540 | 0.94 |
| 469 | XRCC6 | cg19437136 | 0.94 |
| 470 | TBXAS1 | cg01584377 | 0.94 |
| 471 | OR3A2 | cg06841902 | 0.94 |
| 472 | DCPS | cg17631150 | 0.94 |
| 473 | SYNPR | cg13361490 | 0.94 |
| 474 | PAQR6 | cg09604463 | 0.94 |
| 475 | ETNPPL | cg00351892 | 0.94 |
| 476 | ZNF718 | cg26581669 | 0.94 |
| 477 | ELF1 | cg24467989 | 0.94 |
| 478 | RNF222 | cg10218000 | 0.94 |
| 479 | SPC25 | cg00355755 | 0.94 |
| 480 | HSPA12B | cg21700511 | 0.94 |
| 481 | TJP3 | cg19549729 | 0.94 |
| 482 | UHRF1 | cg01369725 | 0.94 |
| 483 | CUL4A | cg10012867 | 0.94 |
| 484 | CLEC12B | cg09565688 | 0.94 |
| 485 | GMPPA | cg03267697 | 0.94 |
| 486 | C20orf111 | cg13681142 | 0.94 |
| 487 | SRC | cg14331199 | 0.94 |
| 488 | HRH4 | cg08043097 | 0.94 |
| 489 | NMNAT1 | cg15156391 | 0.94 |
| 490 | STX1B | cg05787209 | 0.94 |
| 491 | C3orf74 | cg06158985 | 0.94 |
| 492 | THBS2 | cg05969567 | 0.94 |
| 493 | WDR74 | cg14347516 | 0.94 |
| 494 | BAT3 | cg13770361 | 0.93 |
| 495 | SGMS1 | cg21852208 | 0.93 |
| 496 | SCARA3 | cg13351161 | 0.93 |
| 497 | ZNF592 | cg18231690 | 0.93 |
| 498 | C8orf88 | cg20819878 | 0.93 |
| 499 | LOC339539 | cg09941597 | 0.93 |
| 500 | BMP10 | cg10976975 | 0.93 |
| 501 | TRIM26 | cg03782662 | 0.93 |
| 502 | LAIR2 | cg19547155 | 0.93 |
| 503 | FRMD6 | cg22773376 | 0.93 |
| 504 | LOC100129697 | cg16686594 | 0.93 |
| 505 | LYN | cg05973028 | 0.93 |
| 506 | GRPEL1 | cg19408118 | 0.93 |
| 507 | PPT2 | cg20328456 | 0.93 |
| 508 | RG9MTD1 | cg03584693 | 0.93 |
| 509 | PSTPIP2 | cg20468868 | 0.93 |
| 510 | KIF24 | cg14490447 | 0.93 |
| 511 | STAT1 | cg22833204 | 0.93 |
| 512 | MIER2 | cg08617160 | 0.93 |
| 513 | CTDSP2 | cg23105471 | 0.93 |
| 514 | SEMA3C | cg09225287 | 0.93 |
| 515 | HOXC4 | cg00567703 | 0.93 |
| 516 | PLCE1-AS1 | cg15160630 | 0.93 |
| 517 | KIF12 | cg08666741 | 0.93 |
| 518 | RNMT | cg01277202 | 0.93 |
| 519 | GOLSYN | cg07703654 | 0.93 |
| 520 | TEAD1 | cg21709793 | 0.93 |
| 521 | POLR2C | cg04976151 | 0.93 |
| 522 | H19 | cg17769238 | 0.93 |
| 523 | JUP | cg17691598 | 0.93 |
| 524 | FARP2 | cg09639679 | 0.93 |
| 525 | FBXO34 | cg14053764 | 0.93 |
| 526 | MPDU1 | cg13546736 | 0.93 |
| 527 | ALDH1A2 | cg22903883 | 0.93 |
| 528 | PPFIBP2 | cg16964082 | 0.93 |
| 529 | AHCTF1 | cg07507951 | 0.92 |
| 530 | S100A12 | cg05454027 | 0.92 |
| 531 | TNPO2 | cg06347044 | 0.92 |
| 532 | ADCK5 | cg04548483 | 0.92 |
| 533 | PHLDB3 | cg17337072 | 0.92 |
| 534 | PHACTR1 | cg06874640 | 0.92 |
| 535 | SSX2IP | cg18617728 | 0.92 |
| 536 | CTB-178M22.2 | cg24444035 | 0.92 |
| 537 | TMEM14B | cg08662002 | 0.92 |
| 538 | OTOP3 | cg10338787 | 0.92 |
| 539 | RGL1 | cg24706980 | 0.92 |
| 540 | BANP | cg03767934 | 0.92 |
| 541 | CILP2 | cg07002447 | 0.92 |
| 542 | NSMCE1 | cg24052613 | 0.92 |
| 543 | LINC00354 | cg17291325 | 0.92 |
| 544 | TTLL4 | cg01717376 | 0.92 |
| 545 | LPAR5 | cg17874802 | 0.92 |
| 546 | ATP6V1E2 | cg05168133 | 0.92 |
| 547 | NCOA2 | cg20282780 | 0.92 |
| 548 | MAPK8IP2 | cg10341513 | 0.92 |
| 549 | CAMTA2 | cg19427746 | 0.92 |
| 550 | EPHB1 | cg05129878 | 0.92 |
| 551 | MIR449B | cg01988325 | 0.92 |
| 552 | SSB | cg23497644 | 0.92 |
| 553 | HSPBP1 | cg01858556 | 0.92 |
| 554 | DCDC2 | cg16427109 | 0.92 |
| 555 | CHP | cg26299148 | 0.92 |
| 556 | DHRS4 | cg18981338 | 0.92 |
| 557 | COL9A3 | cg15505615 | 0.92 |
| 558 | ZNF331 | cg20646939 | 0.92 |
| 559 | DNASE1L2 | cg18514720 | 0.92 |
| 560 | SYBU | cg08076877 | 0.92 |
| 561 | STK36 | cg11797283 | 0.92 |
| 562 | GORASP2 | cg15617548 | 0.92 |
| 563 | S100Z | cg23745288 | 0.91 |
| 564 | SLC38A1 | cg23031196 | 0.91 |
| 565 | LHFPL2 | cg04357965 | 0.91 |
| 566 | NTSR2 | cg11855727 | 0.91 |
| 567 | ENDOD1 | cg25433915 | 0.91 |
| 568 | DKK4 | cg01762581 | 0.91 |
| 569 | DLD | cg08399831 | 0.91 |
| 570 | MTF1 | cg19823280 | 0.91 |
| 571 | PIPOX | cg23047236 | 0.91 |
| 572 | RXFP4 | cg08403419 | 0.91 |
| 573 | COA6 | cg11155604 | 0.91 |
| 574 | SCNN1B | cg25440020 | 0.91 |
| 575 | HSPA2 | cg26421140 | 0.91 |
| 576 | MAPK10 | cg02898821 | 0.91 |
| 577 | CRY2 | cg15812322 | 0.91 |
| 578 | PIGU | cg05054831 | 0.91 |
| 579 | CDK2AP2 | cg26026771 | 0.91 |
| 580 | FAM107B | cg18124377 | 0.91 |
| 581 | SYT2 | cg05715649 | 0.91 |
| 582 | TMPRSS11F | cg04104541 | 0.91 |
| 583 | RGS3 | cg17066494 | 0.91 |
| 584 | FURIN | cg05466844 | 0.91 |
| 585 | IL1A | cg04859800 | 0.91 |
| 586 | RNPEPL1 | cg07119001 | 0.91 |
| 587 | OSR1 | cg06279113 | 0.91 |
| 588 | C6orf168 | cg00210210 | 0.91 |
| 589 | FLT3LG | cg16891376 | 0.91 |
| 590 | SIGLEC5 | cg27347523 | 0.91 |
| 591 | RBL2 | cg00000029 | 0.91 |
| 592 | DTWD1 | cg07195119 | 0.91 |
| 593 | C1orf65 | cg22372182 | 0.91 |
| 594 | NCOR2 | cg01936957 | 0.91 |
| 595 | BCL7C | cg22377963 | 0.91 |
| 596 | KCNMB1 | cg22646937 | 0.91 |
| 597 | FGF8 | cg22239593 | 0.91 |
| 598 | SERTAD3 | cg26994859 | 0.91 |
| 599 | EMC4 | cg17618596 | 0.91 |
| 600 | TBX10 | cg22385463 | 0.91 |
| 601 | ARHGEF10 | cg27564715 | 0.91 |
| 602 | C18orf1 | cg01041239 | 0.91 |
| 603 | LTA | cg01157951 | 0.91 |
| 604 | FAM193B | cg19658046 | 0.91 |
| 605 | SENP2 | cg07540960 | 0.9 |
| 606 | RASL10A | cg08773625 | 0.9 |
| 607 | ZBTB47 | cg04634182 | 0.9 |
| 608 | ZNF80 | cg22770756 | 0.9 |
| 609 | F7 | cg20983338 | 0.9 |
| 610 | ZNF488 | cg12450316 | 0.9 |
| 611 | RAB37 | cg02834747 | 0.9 |
| 612 | IPO8 | cg17259790 | 0.9 |
| 613 | ALS2CR11 | cg15780361 | 0.9 |
| 614 | RGL4 | cg21444731 | 0.9 |
| 615 | UROC1 | cg05620036 | 0.9 |
| 616 | PITPNM2 | cg22594773 | 0.9 |
| 617 | EP400 | cg08497913 | 0.9 |
| 618 | TP53BP2 | cg13094652 | 0.9 |
| 619 | MORC2-AS1 | cg02256650 | 0.9 |
| 620 | CHST15 | cg24069444 | 0.9 |
| 621 | SLC8B1 | cg25740883 | 0.9 |
| 622 | TBX15 | cg11391335 | 0.9 |
| 623 | ZNF44 | cg26560367 | 0.9 |
| 624 | PLAC8L1 | cg01904727 | 0.9 |
| 625 | UCK1 | cg14352658 | 0.9 |
| 626 | ATP2C1 | cg24097258 | 0.9 |
| 627 | SYCN | cg17574739 | 0.9 |
| 628 | STON1-GTF2A1L | cg19217788 | 0.9 |
| 629 | COL8A2 | cg00358200 | 0.9 |
| 630 | PHF7 | cg10281728 | 0.9 |
| 631 | NDUFS2 | cg01643203 | 0.9 |
| 632 | LINC00368 | cg15987695 | 0.9 |
| 633 | DIRAS1 | cg13363072 | 0.9 |
| 634 | SGK1 | cg09404376 | 0.9 |
| 635 | NFYC | cg20711480 | 0.9 |
| 636 | PRR4 | cg25106783 | 0.9 |
| 637 | TSPAN12 | cg17520932 | 0.9 |
| 638 | RSPO2 | cg12922129 | 0.9 |
| 639 | FOXD2 | cg15868302 | 0.9 |
| 640 | SFMBT1 | cg26850469 | 0.9 |
| 641 | LAT2 | cg03375002 | 0.9 |
| 642 | NTSR2 | cg19584871 | 0.9 |
| 643 | GAB4 | cg19091146 | 0.9 |
| 644 | HYAL2 | cg26460678 | 0.9 |
| 645 | CST5 | cg16365427 | 0.9 |
| 646 | MC2R | cg00335286 | 0.9 |
| 647 | ERICH6-AS1 | cg09281528 | 0.9 |
| 648 | PDE5A | cg10171988 | 0.9 |
| 649 | LRRN4 | cg12453539 | 0.89 |
| 650 | ARHGEF19 | cg05764238 | 0.89 |
| 651 | PCDHB12 | cg17007628 | 0.89 |
| 652 | TRPC6 | cg07931431 | 0.89 |
| 653 | SPATA16 | cg26147262 | 0.89 |
| 654 | TTLL10 | cg14709479 | 0.89 |
| 655 | KCNE1 | cg01183122 | 0.89 |
| 656 | MTMR11 | cg26645700 | 0.89 |
| 657 | TMEM126B | cg21787323 | 0.89 |
| 658 | CPZ | cg04003903 | 0.89 |
| 659 | PLXNA4 | cg24725079 | 0.89 |
| 660 | KCNA3 | cg07903677 | 0.89 |
| 661 | NRXN2 | cg03998212 | 0.89 |
| 662 | C15orf27 | cg25474649 | 0.89 |
| 663 | PEAK1 | cg10192449 | 0.89 |
| 664 | CYB5D2 | cg19105961 | 0.89 |
| 665 | FXR1 | cg16982824 | 0.89 |
| 666 | SMPDL3A | cg23846344 | 0.89 |
| 667 | CRISP2 | cg12440062 | 0.89 |
| 668 | CLIP4 | cg05075112 | 0.89 |
| 669 | PHLDB3 | cg15664905 | 0.89 |
| 670 | KLRG1 | cg00443307 | 0.89 |
| 671 | MYT1L | cg20247911 | 0.89 |
| 672 | NECAP2 | cg13864749 | 0.89 |
| 673 | TAOK3 | cg10833215 | 0.89 |
| 674 | MYOM2 | cg12284098 | 0.89 |
| 675 | BTBD7 | cg09259753 | 0.89 |
| 676 | DIRAS1 | cg11861731 | 0.89 |
| 677 | GNAS | cg00822539 | 0.89 |
| 678 | CLEC2D | cg15060330 | 0.89 |
| 679 | KIAA1644 | cg11099027 | 0.89 |
| 680 | SPATA6L | cg14021777 | 0.89 |
| 681 | LDLRAD3 | cg15208139 | 0.88 |
| 682 | HS3ST3A1 | cg00203913 | 0.88 |
| 683 | SPHKAP | cg00423153 | 0.88 |
| 684 | XAF1 | cg14321269 | 0.88 |
| 685 | TSKU | cg07875873 | 0.88 |
| 686 | SEMA4D | cg13800130 | 0.88 |
| 687 | CBFA2T3 | cg09910271 | 0.88 |
| 688 | LOXL4 | cg04027111 | 0.88 |
| 689 | PPP2R5A | cg11333117 | 0.88 |
| 690 | ZNF721 | cg26262152 | 0.88 |
| 691 | LOC152225 | cg15666147 | 0.88 |
| 692 | KCNQ3 | cg20204316 | 0.88 |
| 693 | SLC35E4 | cg22146903 | 0.88 |
| 694 | NDRG3 | cg11861654 | 0.88 |
| 695 | NPEPL1 | cg01371631 | 0.88 |
| 696 | DLGAP1 | cg16390346 | 0.88 |
| 697 | LRRC2 | cg05226558 | 0.88 |
| 698 | EPS15 | cg01970336 | 0.88 |
| 699 | TMEM17 | cg00301064 | 0.88 |
| 700 | GNG7 | cg06967696 | 0.88 |
| 701 | ZAP70 | cg21773162 | 0.88 |
| 702 | C20orf62 | cg13263208 | 0.88 |
| 703 | CCDC129 | cg13307923 | 0.88 |
| 704 | C11orf24 | cg03007659 | 0.88 |
| 705 | C10orf82 | cg15398400 | 0.88 |
| 706 | USP29 | cg20700952 | 0.88 |
| 707 | GOLT1A | cg20510272 | 0.88 |
| 708 | IKBKE | cg09213696 | 0.88 |
| 709 | CHST8 | cg05746134 | 0.88 |
| 710 | NAP1L4 | cg06538881 | 0.88 |
| 711 | DCTN6 | cg04374813 | 0.88 |
| 712 | GDF10 | cg14588178 | 0.88 |
| 713 | CTSK | cg20802392 | 0.88 |
| 714 | EPN2 | cg08782194 | 0.88 |
| 715 | KIF1C | cg04726697 | 0.88 |
| 716 | STAT5B | cg19465320 | 0.88 |
| 717 | MECOM | cg20202037 | 0.88 |
| 718 | GATSL3 | cg03125765 | 0.88 |
| 719 | GNB1 | cg09535746 | 0.88 |
| 720 | CATSPER4 | cg15910554 | 0.88 |
| 721 | GPX3 | cg01378878 | 0.88 |
| 722 | MIR181C | cg08310363 | 0.88 |
| 723 | IL17C | cg04810063 | 0.88 |
| 724 | KCNJ8 | cg22083892 | 0.88 |
| 725 | C3orf74 | cg24277586 | 0.88 |
| 726 | PNMAL2 | cg01029669 | 0.88 |
| 727 | ZNF778 | cg22050642 | 0.88 |
| 728 | DMKN | cg16677448 | 0.88 |
| 729 | TAS2R1 | cg18027004 | 0.88 |
| 730 | STARD3 | cg26161820 | 0.88 |
| 731 | SLC2A8 | cg06843131 | 0.87 |
| 732 | CDC14C | cg16207528 | 0.87 |
| 733 | MEDAG | cg25226285 | 0.87 |
| 734 | GULP1 | cg07077227 | 0.87 |
| 735 | DGCR8 | cg18757169 | 0.87 |
| 736 | RUNX3 | cg05714960 | 0.87 |
| 737 | RARA | cg25362050 | 0.87 |
| 738 | TRIP10 | cg02085507 | 0.87 |
| 739 | KRTAP20-2 | cg00948500 | 0.87 |
| 740 | C20orf4 | cg23506077 | 0.87 |
| 741 | OR2T33 | cg06400301 | 0.87 |
| 742 | KANSL1 | cg02182504 | 0.87 |
| 743 | PLAGL1 | cg14161241 | 0.87 |
| 744 | CKM | cg07165026 | 0.87 |
| 745 | BBX | cg02824953 | 0.87 |
| 746 | TMEM82 | cg20843478 | 0.87 |
| 747 | CD59 | cg03292992 | 0.87 |
| 748 | RBMXL2 | cg23916104 | 0.87 |
| 749 | INPP4B | cg14287071 | 0.87 |
| 750 | SSTR4 | cg14631053 | 0.87 |
| 751 | CUEDC1 | cg13830856 | 0.87 |
| 752 | COL1A2 | cg24406898 | 0.87 |
| 753 | SPEF1 | cg08708079 | 0.87 |
| 754 | LOC650226 | cg17328716 | 0.87 |
| 755 | KLK5 | cg27583102 | 0.87 |
| 756 | GPN3 | cg19666600 | 0.87 |
| 757 | DKFZp434L192 | cg17876758 | 0.87 |
| 758 | SMTNL1 | cg23362108 | 0.87 |
| 759 | PARD6G | cg16553796 | 0.87 |
| 760 | S100A12 | cg15108991 | 0.87 |
| 761 | H2AFY | cg13021507 | 0.87 |
| 762 | INSIG1 | cg27127927 | 0.87 |
| 763 | SLC6A1-AS1 | cg04075866 | 0.87 |
| 764 | TTBK1 | cg10203631 | 0.87 |
| 765 | RLN3 | cg18256471 | 0.87 |
| 766 | MUC15 | cg07956154 | 0.87 |
| 767 | LOC100507487 | cg26013916 | 0.87 |
| 768 | LINC00411 | cg05219813 | 0.87 |
| 769 | NECAB1 | cg00151250 | 0.87 |
| 770 | CHGA | cg04453367 | 0.87 |
| 771 | PLA2G2A | cg02520499 | 0.87 |
| 772 | CACNA1C-IT2 | cg01260909 | 0.87 |
| 773 | FAM19A1 | cg07192748 | 0.87 |
| 774 | C10orf18 | cg09411597 | 0.87 |
| 775 | BHLHA15 | cg22042803 | 0.87 |
| 776 | JUP | cg00666845 | 0.87 |
| 777 | TSHZ1 | cg13336350 | 0.87 |
| 778 | COPZ1 | cg02507455 | 0.87 |
| 779 | C15orf48 | cg11429271 | 0.87 |
| 780 | PLS1 | cg16488116 | 0.87 |
| 781 | CXCL12 | cg11267527 | 0.86 |
| 782 | PRKCA | cg09645572 | 0.86 |
| 783 | PGCP | cg01892689 | 0.86 |
| 784 | EEF1B2 | cg04202957 | 0.86 |
| 785 | LYPD5 | cg11993925 | 0.86 |
| 786 | DENND2D | cg19268695 | 0.86 |
| 787 | SQSTM1 | cg09518506 | 0.86 |
| 788 | CPT1A | cg22911054 | 0.86 |
| 789 | NEDD4L | cg16576811 | 0.86 |
| 790 | ACP1 | cg26398848 | 0.86 |
| 791 | FAM159A | cg26845363 | 0.86 |
| 792 | IL16 | cg12566412 | 0.86 |
| 793 | PPIF | cg17037153 | 0.86 |
| 794 | NDST1 | cg14873515 | 0.86 |
| 795 | TP73 | cg04137576 | 0.86 |
| 796 | TMEM210 | cg05484618 | 0.86 |
| 797 | INVS | cg25241040 | 0.86 |
| 798 | ADAMTSL5 | cg23572813 | 0.86 |
| 799 | ST3GAL1 | cg01024504 | 0.86 |
| 800 | SLC6A12 | cg07904051 | 0.86 |
| 801 | EPHA3 | cg16797972 | 0.86 |
| 802 | AP2M1 | cg20586996 | 0.86 |
| 803 | CLK2 | cg03212674 | 0.86 |
| 804 | SIPA1L3 | cg09318848 | 0.86 |
| 805 | RAPGEF4 | cg17229453 | 0.86 |
| 806 | TMEM203 | cg16153652 | 0.86 |
| 807 | TCP11L2 | cg06955856 | 0.86 |
| 808 | BHLHA9 | cg24605747 | 0.86 |
| 809 | CHRNA1 | cg04859027 | 0.86 |
| 810 | SNORD116-12 | cg03411305 | 0.86 |
| 811 | MAGI2 | cg24391460 | 0.86 |
| 812 | RIMS3 | cg16359657 | 0.86 |
| 813 | NBN | cg14204925 | 0.86 |
| 814 | PELP1 | cg23921860 | 0.86 |
| 815 | TMEM219 | cg03886752 | 0.86 |
| 816 | HFM1 | cg09737265 | 0.86 |
| 817 | GDA | cg17395154 | 0.86 |
| 818 | PRR23C | cg06508738 | 0.86 |
| 819 | CHRM2 | cg26253500 | 0.86 |
| 820 | ZNF324B | cg17725398 | 0.86 |
| 821 | L3MBTL3 | cg20719116 | 0.86 |
| 822 | CCDC65 | cg19978583 | 0.86 |
| 823 | AVP | cg05136169 | 0.85 |
| 824 | CASC10 | cg23366556 | 0.85 |
| 825 | TMEM93 | cg13345380 | 0.85 |
| 826 | RNF207 | cg19180828 | 0.85 |
| 827 | RAB7A | cg13176762 | 0.85 |
| 828 | LINC00371 | cg04516854 | 0.85 |
| 829 | LRRN4 | cg20918682 | 0.85 |
| 830 | SEC14L6 | cg15125707 | 0.85 |
| 831 | UBLCP1 | cg02884181 | 0.85 |
| 832 | CTTNBP2NL | cg26875637 | 0.85 |
| 833 | KRTAP10-2 | cg24322131 | 0.85 |
| 834 | ZBED3 | cg05278955 | 0.85 |
| 835 | DKFZP434H168 | cg04507426 | 0.85 |
| 836 | ZNF284 | cg14674574 | 0.85 |
| 837 | PTPRT | cg10348362 | 0.85 |
| 838 | FANCI | cg08433293 | 0.85 |
| 839 | SNORA38 | cg07708721 | 0.85 |
| 840 | INHBC | cg08640498 | 0.85 |
| 841 | SNORD116-1 | cg04061361 | 0.85 |
| 842 | P4HA2 | cg24117468 | 0.85 |
| 843 | SMPDL3B | cg05320933 | 0.85 |
| 844 | AKR7A2 | cg13494428 | 0.85 |
| 845 | KSR1 | cg17585031 | 0.85 |
| 846 | LOC101927822 | cg15550804 | 0.85 |
| 847 | NR3C1 | cg18718518 | 0.85 |
| 848 | FAM19A1 | cg01709518 | 0.85 |
| 849 | LOC286238 | cg03578548 | 0.85 |
| 850 | WSCD2 | cg03626024 | 0.85 |
| 851 | SLC23A2 | cg16348812 | 0.85 |
| 852 | RCOR2 | cg24829313 | 0.85 |
| 853 | DOK3 | cg07265015 | 0.85 |
| 854 | LEFTY2 | cg20742696 | 0.85 |
| 855 | ACOX2 | cg13705284 | 0.85 |
| 856 | CD79A | cg19825589 | 0.85 |
| 857 | LDLRAD4 | cg07195078 | 0.85 |
| 858 | SLC22A23 | cg13415859 | 0.85 |
| 859 | POU2F2 | cg07667560 | 0.85 |
| 860 | SNRPN | cg25657700 | 0.85 |
| 861 | ZNF106 | cg16128973 | 0.85 |
| 862 | CRNKL1 | cg04507075 | 0.85 |
| 863 | NEUROG1 | cg24899138 | 0.85 |
| 864 | IL32 | cg23813257 | 0.85 |
| 865 | LOC338797 | cg22317906 | 0.85 |
| 866 | PPP1R10 | cg04862321 | 0.85 |
| 867 | SVIL | cg00622375 | 0.85 |
| 868 | GKAP1 | cg17375598 | 0.85 |
| 869 | C18orf21 | cg12580471 | 0.85 |
| 870 | GREB1 | cg25472172 | 0.85 |
| 871 | LOC100189589 | cg00043800 | 0.85 |
| 872 | MIR6889 | cg04424420 | 0.85 |
| 873 | CLEC4GP1 | cg23190402 | 0.84 |
| 874 | PDE6B | cg01152688 | 0.84 |
| 875 | SNORD115-32 | cg15153347 | 0.84 |
| 876 | SEPT9 | cg12424499 | 0.84 |
| 877 | NIM1K | cg23013372 | 0.84 |
| 878 | KCNA4 | cg20213416 | 0.84 |
| 879 | HOXA4 | cg08657492 | 0.84 |
| 880 | APOC1P1 | cg08121984 | 0.84 |
| 881 | NGF | cg04082532 | 0.84 |
| 882 | TTBK1 | cg25521481 | 0.84 |
| 883 | LOC388780 | cg21498923 | 0.84 |
| 884 | LIMS1 | cg18232975 | 0.84 |
| 885 | SLC37A2 | cg11537406 | 0.84 |
| 886 | ANXA11 | cg02711637 | 0.84 |
| 887 | BIRC7 | cg25099532 | 0.84 |
| 888 | C6orf47 | cg18842363 | 0.84 |
| 889 | TRAF5 | cg12799170 | 0.84 |
| 890 | TLK1 | cg08863939 | 0.84 |
| 891 | SYT3 | cg01775986 | 0.84 |
| 892 | TM4SF19-AS1 | cg12241552 | 0.84 |
| 893 | CD3D | cg03074244 | 0.84 |
| 894 | PACSIN2 | cg05330000 | 0.84 |
| 895 | LINC01072 | cg21526187 | 0.84 |
| 896 | CREB3L1 | cg12256550 | 0.84 |
| 897 | HIVEP2 | cg04356230 | 0.84 |
| 898 | SGMS1 | cg11500842 | 0.84 |
| 899 | BEND4 | cg14632941 | 0.84 |
| 900 | C22orf32 | cg18034295 | 0.84 |
| 901 | SYCP1 | cg24985030 | 0.84 |
| 902 | SRMS | cg17793372 | 0.84 |
| 903 | HMBS | cg16169193 | 0.84 |
| 904 | EMP1 | cg21490635 | 0.84 |
| 905 | NACC2 | cg11598034 | 0.84 |
| 906 | KCTD1 | cg22277092 | 0.84 |
| 907 | GRAMD2 | cg19687230 | 0.84 |
| 908 | ST6GAL1 | cg11386711 | 0.84 |
| 909 | DACT3 | cg16262525 | 0.84 |
| 910 | C10orf96 | cg13836550 | 0.84 |
| 911 | FAM184B | cg00306311 | 0.84 |
| 912 | KRTAP10-11 | cg18605642 | 0.84 |
| 913 | EPS8L1 | cg24402300 | 0.84 |
| 914 | EDARADD | cg09809672 | 0.84 |
| 915 | ITGA2B | cg03609058 | 0.84 |
| 916 | NDUFA7 | cg03633120 | 0.84 |
| 917 | NFATC2IP | cg22745280 | 0.84 |
| 918 | SYNDIG1L | cg15713972 | 0.84 |
| 919 | TMEM72 | cg05187965 | 0.84 |
| 920 | KCNK3 | cg06313935 | 0.84 |
| 921 | LAMC3 | cg07725394 | 0.84 |
| 922 | CORO2B | cg15813838 | 0.84 |
| 923 | CTDP1 | cg13598593 | 0.84 |
| 924 | ZNF683 | cg15118519 | 0.84 |
| 925 | DEFB125 | cg14814714 | 0.84 |
| 926 | GGT1 | cg13183251 | 0.84 |
| 927 | TRPM8 | cg08629502 | 0.84 |
| 928 | KRTAP13-3 | cg12141967 | 0.83 |
| 929 | OBSL1 | cg19780389 | 0.83 |
| 930 | SEZ6 | cg19929340 | 0.83 |
| 931 | PPFIBP1 | cg27272447 | 0.83 |
| 932 | TCL1B | cg05402891 | 0.83 |
| 933 | LYST | cg16962115 | 0.83 |
| 934 | KCNA6 | cg22087651 | 0.83 |
| 935 | LOC100292680 | cg18071202 | 0.83 |
| 936 | TPCN2 | cg23247191 | 0.83 |
| 937 | KLK3 | cg08466913 | 0.83 |
| 938 | STK19 | cg13069441 | 0.83 |
| 939 | C16orf82 | cg17306401 | 0.83 |
| 940 | VANGL1 | cg25773695 | 0.83 |
| 941 | LRRC38 | cg27044429 | 0.83 |
| 942 | USP2 | cg04125136 | 0.83 |
| 943 | IL15 | cg26867774 | 0.83 |
| 944 | DAB1 | cg06710648 | 0.83 |
| 945 | SERINC5 | cg15181688 | 0.83 |
| 946 | TMEM246-AS1 | cg07605566 | 0.83 |
| 947 | IL9 | cg12575771 | 0.83 |
| 948 | ODF3L1 | cg18524420 | 0.83 |
| 949 | SLC39A1 | cg08088996 | 0.83 |
| 950 | ATP2B4 | cg19087104 | 0.83 |
| 951 | SGCE | cg21771834 | 0.83 |
| 952 | CASP7 | cg09500881 | 0.83 |
| 953 | ZNF790 | cg26095210 | 0.83 |
| 954 | HSD3B7 | cg05539847 | 0.83 |
| 955 | DBR1 | cg10065081 | 0.83 |
| 956 | ZNF304 | cg03699054 | 0.83 |
| 957 | TMEM133 | cg06965373 | 0.83 |
| 958 | CCDC60 | cg27435644 | 0.83 |
| 959 | TTC9 | cg03762817 | 0.83 |
| 960 | TBXAS1 | cg06904057 | 0.83 |
| 961 | MIR1249 | cg04880355 | 0.83 |
| 962 | TNFRSF8 | cg01890404 | 0.83 |
| 963 | DMGDH | cg01856645 | 0.83 |
| 964 | MATN4 | cg14608505 | 0.83 |
| 965 | LOC101927464 | cg11490836 | 0.83 |
| 966 | CASP7 | cg10572337 | 0.83 |
| 967 | SLC12A8 | cg14391622 | 0.83 |
| 968 | CCNB1IP1 | cg18728631 | 0.83 |
| 969 | ABHD12B | cg13335112 | 0.82 |
| 970 | LOC102723544 | cg20921778 | 0.82 |
| 971 | GCC1 | cg08586737 | 0.82 |
| 972 | TMCO4 | cg20993090 | 0.82 |
| 973 | IGFBP5 | cg22522886 | 0.82 |
| 974 | COL9A2 | cg04100190 | 0.82 |
| 975 | MAP2K6 | cg04098023 | 0.82 |
| 976 | IGF2 | cg01921126 | 0.82 |
| 977 | CLN8 | cg14813353 | 0.82 |
| 978 | AUH | cg03254601 | 0.82 |
| 979 | GOLSYN | cg02389590 | 0.82 |
| 980 | F13A1 | cg11506733 | 0.82 |
| 981 | SRRM3 | cg01475729 | 0.82 |
| 982 | NCOA1 | cg14337036 | 0.82 |
| 983 | CHD7 | cg05575639 | 0.82 |
| 984 | OLFM1 | cg08268099 | 0.82 |
| 985 | ANGPTL2 | cg14281592 | 0.82 |
| 986 | NT5C2 | cg20912449 | 0.82 |
| 987 | INPP5J | cg24810654 | 0.82 |
| 988 | FLJ39609 | cg19159092 | 0.82 |
| 989 | CELF6 | cg02935847 | 0.82 |
| 990 | DLGAP2 | cg17822710 | 0.82 |
| 991 | GCA | cg03452895 | 0.82 |
| 992 | SLC6A19 | cg26711638 | 0.82 |
| 993 | CPZ | cg23283187 | 0.82 |
| 994 | PDK1 | cg10165864 | 0.82 |
| 995 | RP1L1 | cg02030396 | 0.82 |
| 996 | MYOM3 | cg06649280 | 0.82 |
| 997 | EMILIN1 | cg19399165 | 0.82 |
| 998 | C17orf75 | cg18643150 | 0.82 |
| 999 | ZNF205 | cg10899887 | 0.82 |
| 1000 | TMEM215 | cg06600697 | 0.82 |
| 1001 | ANKRD17 | cg00539511 | 0.82 |
| 1002 | TMEM105 | cg01905773 | 0.82 |
| 1003 | KCTD13 | cg02747254 | 0.82 |
| 1004 | MNDA | cg16663980 | 0.82 |
| 1005 | SV2B | cg25969814 | 0.82 |
| 1006 | TCTN2 | cg05384724 | 0.82 |
| 1007 | ZFAT | cg14953610 | 0.82 |
| 1008 | CD276 | cg10586317 | 0.82 |
| 1009 | OXT | cg07597882 | 0.82 |
| 1010 | PYGB | cg14384714 | 0.82 |
| 1011 | TEC | cg13294560 | 0.82 |
| 1012 | OR5D16 | cg00899834 | 0.82 |
| 1013 | SLC23A3 | cg18822719 | 0.82 |
| 1014 | RBFOX3 | cg16284924 | 0.81 |
| 1015 | LDLRAD4 | cg18033728 | 0.81 |
| 1016 | ZBTB16 | cg11031278 | 0.81 |
| 1017 | GREB1 | cg11534680 | 0.81 |
| 1018 | RAMP1 | cg24887490 | 0.81 |
| 1019 | C11orf86 | cg07962796 | 0.81 |
| 1020 | BMPR1A | cg18525126 | 0.81 |
| 1021 | NPLOC4 | cg12978798 | 0.81 |
| 1022 | AKNA | cg04740744 | 0.81 |
| 1023 | CES3 | cg11834064 | 0.81 |
| 1024 | BBX | cg15916331 | 0.81 |
| 1025 | SRP9 | cg21188007 | 0.81 |
| 1026 | SH3BP2 | cg22198044 | 0.81 |
| 1027 | TBXAS1 | cg25287523 | 0.81 |
| 1028 | APOL4 | cg11420782 | 0.81 |
| 1029 | KCNH2 | cg14711997 | 0.81 |
| 1030 | DCN | cg02322911 | 0.81 |
| 1031 | TDRD7 | cg03446675 | 0.81 |
| 1032 | ZNF276 | cg02252742 | 0.81 |
| 1033 | ANXA6 | cg04150429 | 0.81 |
| 1034 | SPIRE1 | cg11337929 | 0.81 |
| 1035 | CASZ1 | cg00787856 | 0.81 |
| 1036 | CCNB1 | cg25474616 | 0.81 |
| 1037 | S100A10 | cg07908047 | 0.81 |
| 1038 | HORMAD2 | cg16686158 | 0.81 |
| 1039 | MIR194-2 | cg06104877 | 0.81 |
| 1040 | ZBTB7C | cg16078269 | 0.81 |
| 1041 | LASS2 | cg07422880 | 0.81 |
| 1042 | RHBG | cg17935233 | 0.81 |
| 1043 | GIPR | cg00500037 | 0.81 |
| 1044 | CEBPD | cg25661026 | 0.81 |
| 1045 | NLRP11 | cg03789934 | 0.81 |
| 1046 | C11orf41 | cg11623855 | 0.81 |
| 1047 | FBXO16 | cg16145757 | 0.81 |
| 1048 | CHSY1 | cg23222772 | 0.81 |
| 1049 | ITGAE | cg13984928 | 0.81 |
| 1050 | LIMCH1 | cg10534544 | 0.81 |
| 1051 | PRDM11 | cg14791029 | 0.81 |
| 1052 | GNG7 | cg23463608 | 0.81 |
| 1053 | TXNL1 | cg24431033 | 0.81 |
| 1054 | SRMS | cg22442730 | 0.81 |
| 1055 | GNAL | cg00576132 | 0.81 |
| 1056 | NXPH2 | cg23302570 | 0.81 |
| 1057 | HDDC3 | cg11145826 | 0.81 |
| 1058 | C10orf58 | cg19862899 | 0.81 |
| 1059 | EPGN | cg07796901 | 0.81 |
| 1060 | SMARCA5-AS1 | cg25195877 | 0.81 |
| 1061 | RPS6KA1 | cg22512670 | 0.81 |
| 1062 | AMIGO3 | cg05666287 | 0.8 |
| 1063 | FRRS1 | cg04308687 | 0.8 |
| 1064 | RAPSN | cg27466532 | 0.8 |
| 1065 | ADAMTSL5 | cg26732521 | 0.8 |
| 1066 | SCOC | cg20424311 | 0.8 |
| 1067 | DMWD | cg16681516 | 0.8 |
| 1068 | DIRAS3 | cg13605615 | 0.8 |
| 1069 | TEAD4 | cg01039401 | 0.8 |
| 1070 | PRKAR1B | cg23605961 | 0.8 |
| 1071 | PCNA | cg09445550 | 0.8 |
| 1072 | EDNRA | cg20402736 | 0.8 |
| 1073 | ITLN1 | cg10094191 | 0.8 |
| 1074 | FLJ44606 | cg05001932 | 0.8 |
| 1075 | ACTR1B | cg05184570 | 0.8 |
| 1076 | LOC148824 | cg12758973 | 0.8 |
| 1077 | PCDHB4 | cg13288164 | 0.8 |
| 1078 | LOC101929577 | cg16662291 | 0.8 |
| 1079 | CTBP2 | cg25961397 | 0.8 |
| 1080 | DNAJB5 | cg14644378 | 0.8 |
| 1081 | PGF | cg19959591 | 0.8 |
| 1082 | IGFBP1 | cg00110785 | 0.8 |
| 1083 | ZMIZ1 | cg24983858 | 0.8 |
| 1084 | RGS14 | cg12377595 | 0.8 |
| 1085 | LOC732275 | cg07118299 | 0.8 |
| 1086 | ATP2B2 | cg17622397 | 0.8 |
| 1087 | MBOAT4 | cg10471143 | 0.8 |
| 1088 | CPA5 | cg12936509 | 0.8 |
| 1089 | ZNF430 | cg23882790 | 0.8 |
| 1090 | CDA | cg05646357 | 0.8 |
| 1091 | TNFSF12 | cg19378473 | 0.8 |
| 1092 | KANK1 | cg02596384 | 0.8 |
| 1093 | TMEM8C | cg01263574 | 0.79 |
| 1094 | LOC100192379 | cg13640145 | 0.79 |
| 1095 | LRRC14B | cg03553576 | 0.79 |
| 1096 | NEUROD1 | cg20806345 | 0.79 |
| 1097 | CCDC148 | cg13896783 | 0.79 |
| 1098 | RFWD2 | cg25623035 | 0.79 |
| 1099 | PACS2 | cg09716042 | 0.79 |
| 1100 | CCDC12 | cg24194630 | 0.79 |
| 1101 | PXN | cg20153322 | 0.79 |
| 1102 | DEPDC6 | cg02919712 | 0.79 |
| 1103 | MCOLN3 | cg12210618 | 0.79 |
| 1104 | PPARG | cg17369845 | 0.79 |
| 1105 | ST7-AS2 | cg09584483 | 0.79 |
| 1106 | TRIML1 | cg25760325 | 0.79 |
| 1107 | DTX3 | cg04492438 | 0.79 |
| 1108 | LINC00950 | cg15337981 | 0.79 |
| 1109 | MRPL21 | cg11792649 | 0.79 |
| 1110 | CD226 | cg13164537 | 0.79 |
| 1111 | CRYAB | cg15545878 | 0.79 |
| 1112 | COMMD7 | cg23356674 | 0.79 |
| 1113 | C11orf94 | cg02164088 | 0.79 |
| 1114 | PABPC4 | cg16510040 | 0.79 |
| 1115 | AMPD3 | cg11325786 | 0.79 |
| 1116 | FKBP5 | cg03591753 | 0.79 |
| 1117 | USP29 | cg16547341 | 0.79 |
| 1118 | CUGBP1 | cg25953688 | 0.79 |
| 1119 | TEX2 | cg00864012 | 0.79 |
| 1120 | MS4A10 | cg11633310 | 0.79 |
| 1121 | ENTPD3-AS1 | cg07018561 | 0.79 |
| 1122 | BRI3BP | cg22867594 | 0.79 |
| 1123 | LOC286094 | cg13980233 | 0.79 |
| 1124 | PSMF1 | cg20611399 | 0.79 |
| 1125 | NOSIP | cg09823713 | 0.79 |
| 1126 | C16orf93 | cg06591842 | 0.79 |
| 1127 | SIRPA | cg27635450 | 0.79 |
| 1128 | LINC01570 | cg26150969 | 0.79 |
| 1129 | LIMS1 | cg09176441 | 0.79 |
| 1130 | NRIP2 | cg02852959 | 0.79 |
| 1131 | MIR3621 | cg13224090 | 0.79 |
| 1132 | RHBG | cg07702888 | 0.79 |
| 1133 | SLC26A10 | cg08820231 | 0.79 |
| 1134 | KIAA1949 | cg27385757 | 0.79 |
| 1135 | GJB4 | cg07594531 | 0.78 |
| 1136 | GPD2 | cg09290120 | 0.78 |
| 1137 | CCL22 | cg02532928 | 0.78 |
| 1138 | PAK6 | cg01279378 | 0.78 |
| 1139 | NOTCH3 | cg25902639 | 0.78 |
| 1140 | ARHGEF7 | cg25759064 | 0.78 |
| 1141 | C2orf58 | cg05415606 | 0.78 |
| 1142 | C12orf76 | cg01753950 | 0.78 |
| 1143 | C14orf159 | cg05131266 | 0.78 |
| 1144 | LOC100134391 | cg22008124 | 0.78 |
| 1145 | LHFPL4 | cg02530165 | 0.78 |
| 1146 | BMP7 | cg04043642 | 0.78 |
| 1147 | RBM46 | cg05914674 | 0.78 |
| 1148 | GPR77 | cg16734795 | 0.78 |
| 1149 | CPNE6 | cg13711543 | 0.78 |
| 1150 | ACAN | cg26410149 | 0.78 |
| 1151 | RIMS4 | cg21656205 | 0.78 |
| 1152 | SHANK2 | cg07913800 | 0.78 |
| 1153 | NRXN2 | cg01475205 | 0.78 |
| 1154 | OSBPL2 | cg26689913 | 0.78 |
| 1155 | ESRRG | cg04217177 | 0.78 |
| 1156 | CKAP5 | cg16136385 | 0.77 |
| 1157 | PCDHB17 | cg04056384 | 0.77 |
| 1158 | GNG12 | cg05040656 | 0.77 |
| 1159 | LOC134466 | cg08747963 | 0.77 |
| 1160 | OR10T2 | cg24982343 | 0.77 |
| 1161 | FAM90A1 | cg20633835 | 0.77 |
| 1162 | RTEL1 | cg02062530 | 0.77 |
| 1163 | BANP | cg20611109 | 0.77 |
| 1164 | KCNJ10 | cg10654598 | 0.77 |
| 1165 | LOC100506388 | cg21740158 | 0.77 |
| 1166 | C9orf103 | cg14512326 | 0.77 |
| 1167 | CORO1C | cg10667794 | 0.77 |
| 1168 | ANKRD26P1 | cg02017347 | 0.77 |
| 1169 | NCOR1 | cg08125503 | 0.77 |
| 1170 | NDUFB9 | cg17680872 | 0.77 |
| 1171 | GNG7 | cg22570676 | 0.77 |
| 1172 | VARS2 | cg09424348 | 0.77 |
| 1173 | CACNB2 | cg14094927 | 0.77 |
| 1174 | WRAP53 | cg21840333 | 0.76 |
| 1175 | C6orf154 | cg11523799 | 0.76 |
| 1176 | BHLHA15 | cg20061890 | 0.76 |
| 1177 | C7orf73 | cg07804024 | 0.76 |
| 1178 | ITGA8 | cg07712540 | 0.76 |
| 1179 | SHANK1 | cg14426373 | 0.76 |
| 1180 | BANP | cg01357591 | 0.76 |
| 1181 | EVA1A | cg22510460 | 0.76 |
| 1182 | LIMS1 | cg19752397 | 0.76 |
| 1183 | DICER1 | cg04112704 | 0.76 |
| 1184 | WARS | cg04899753 | 0.76 |
| 1185 | CAPN14 | cg05082258 | 0.76 |
| 1186 | ERMN | cg05081953 | 0.76 |
| 1187 | SUSD1 | cg14038618 | 0.76 |
| 1188 | ERC1 | cg06708720 | 0.76 |
| 1189 | MMS22L | cg19562453 | 0.76 |
| 1190 | ZNF578 | cg03774704 | 0.76 |
| 1191 | ALG2 | cg11148808 | 0.75 |
| 1192 | SATB1 | cg20016068 | 0.75 |
| 1193 | PEG10 | cg27435646 | 0.75 |
| 1194 | CDC14B | cg13226907 | 0.75 |
| 1195 | LRRC33 | cg20534585 | 0.75 |
| 1196 | SORCS1 | cg07392385 | 0.75 |
| 1197 | GULP1 | cg12268101 | 0.75 |
| 1198 | NNMT | cg15560405 | 0.75 |
| 1199 | RPS15 | cg19716018 | 0.75 |
| 1200 | UGT3A1 | cg05512756 | 0.75 |
| 1201 | MIR153-2 | cg10893656 | 0.75 |
| 1202 | KLK4 | cg00329745 | 0.75 |
| 1203 | CNPY1 | cg15983026 | 0.75 |
| 1204 | LOC101929413 | cg09109562 | 0.74 |
| 1205 | TTBK1 | cg17699330 | 0.74 |
| 1206 | TDRD5 | cg09656934 | 0.74 |
| 1207 | RBP1 | cg06726820 | 0.74 |
| 1208 | MAMSTR | cg12234455 | 0.74 |
| 1209 | C11orf85 | cg08697310 | 0.74 |
| 1210 | KIAA1239 | cg06805348 | 0.74 |
| 1211 | MTIF3 | cg20147645 | 0.74 |
| 1212 | PRRC1 | cg20690028 | 0.74 |
| 1213 | COL26A1 | cg25916759 | 0.74 |
| 1214 | LOC100287216 | cg25308803 | 0.74 |
| 1215 | TRIP13 | cg16417200 | 0.74 |
| 1216 | GRM2 | cg21899500 | 0.74 |
| 1217 | CCDC150 | cg01265662 | 0.74 |
| 1218 | CLN8 | cg26975524 | 0.74 |
| 1219 | PRAME | cg19804859 | 0.74 |
| 1220 | CCDC30 | cg24501381 | 0.74 |
| 1221 | PAK7 | cg07439609 | 0.74 |
| 1222 | ALOX15 | cg11334406 | 0.74 |
| 1223 | TUBB3 | cg24272172 | 0.74 |
| 1224 | C1orf35 | cg22790377 | 0.74 |
| 1225 | PKNOX1 | cg18945945 | 0.73 |
| 1226 | C7orf31 | cg02544454 | 0.73 |
| 1227 | CHI3L1 | cg19081101 | 0.73 |
| 1228 | ADD3 | cg05214460 | 0.73 |
| 1229 | FGR | cg00404394 | 0.73 |
| 1230 | TNNT1 | cg21127534 | 0.73 |
| 1231 | C1orf100 | cg09033006 | 0.73 |
| 1232 | MIR4766 | cg09916443 | 0.73 |
| 1233 | PPARG | cg27051533 | 0.73 |
| 1234 | UNC13A | cg25731534 | 0.72 |
| 1235 | ZNF264 | cg27176357 | 0.72 |
| 1236 | RNPEP | cg03981535 | 0.72 |
| 1237 | ITGAE | cg09552070 | 0.72 |
| 1238 | METRNL | cg01908551 | 0.72 |
| 1239 | CLDN1 | cg03597510 | 0.72 |
| 1240 | CCDC80 | cg16198723 | 0.71 |
| 1241 | TNFRSF8 | cg13457900 | 0.71 |
| 1242 | GALNTL2 | cg10453019 | 0.71 |
| 1243 | RASL11B | cg03786743 | 0.71 |
| 1244 | DTNA | cg18420599 | 0.71 |
| 1245 | STX5 | cg07112634 | 0.71 |
| 1246 | GNG4 | cg10866988 | 0.71 |
| 1247 | CETP | cg12564453 | 0.71 |
| 1248 | LOC401010 | cg22656550 | 0.71 |
| 1249 | MUM1 | cg08643131 | 0.71 |
| 1250 | LOC101928731 | cg14229001 | 0.71 |
| 1251 | IP6K3 | cg08280829 | 0.71 |
| 1252 | KCNA3 | cg00995520 | 0.71 |
| 1253 | CTBP1 | cg15586393 | 0.7 |
| 1254 | ALOX15 | cg22249932 | 0.7 |
| 1255 | BBS2 | cg07280206 | 0.7 |
| 1256 | CPXM2 | cg08627010 | 0.7 |
| 1257 | SORD | cg25426733 | 0.7 |
| 1258 | RNF126 | cg03605761 | 0.7 |
| 1259 | PSMG3 | cg10712491 | 0.7 |
| 1260 | CCBL2 | cg11309454 | 0.7 |
| 1261 | ZC3H3 | cg03870862 | 0.69 |
| 1262 | COL2A1 | cg08886711 | 0.69 |
| 1263 | SCAF11 | cg08122691 | 0.69 |
| 1264 | RPH3AL | cg19446165 | 0.69 |
| 1265 | FAM135B | cg26005485 | 0.69 |
| 1266 | ZNF536 | cg23458168 | 0.69 |
| 1267 | TMEM204 | cg16336651 | 0.69 |
| 1268 | GORASP2 | cg14744970 | 0.68 |
| 1269 | GATAD2A | cg13934873 | 0.68 |
| 1270 | UNC79 | cg02027651 | 0.68 |
| 1271 | STMN2 | cg08496775 | 0.68 |
| 1272 | EIF4E3 | cg24848973 | 0.67 |
| 1273 | WSCD2 | cg12713877 | 0.66 |
| 1274 | REEP3 | cg11226562 | 0.66 |
| 1275 | ENOSF1 | cg17732033 | 0.66 |
| 1276 | ST6GAL2 | cg19501858 | 0.66 |
| 1277 | NDRG4 | cg24093474 | 0.66 |
| 1278 | ALOX15 | cg17328062 | 0.66 |
| 1279 | RAPGEF1 | cg05283385 | 0.66 |
| 1280 | PALLD | cg14648920 | 0.65 |
| 1281 | SHANK2 | cg18248145 | 0.65 |
| 1282 | C8orf33 | cg24480555 | 0.65 |
| 1283 | GP6 | cg25818583 | 0.65 |
| 1284 | HOXA5 | cg09549073 | 0.65 |
| 1285 | SLC6A1 | cg17840583 | 0.65 |
| 1286 | HPS5 | cg03298714 | 0.64 |
| 1287 | ZNF454 | cg02031359 | 0.63 |
| 1288 | S100A16 | cg23505766 | 0.63 |
| 1289 | NDRG4 | cg17107388 | 0.61 |
| 1290 | MIR487B | cg02882979 | 0.59 |
| 1291 | FAM110C | cg20318662 | 0.58 |
| 1292 | PIWIL1 | cg19424457 | 0.57 |
| 1293 | ANTXRL | cg10430941 | 0.56 |
| 1294 | HOPX | cg19978674 | 0.56 |
| 1295 | S100A13 | cg06562291 | 0.55 |
| 1296 | DTNB | cg08459290 | 0.55 |
